# Immunogenicity and Safety of PHH-1V COVID-19 Vaccine as a Heterologous Booster in Adolescents

**DOI:** 10.1101/2025.10.08.25337592

**Authors:** Ignasi Esteban Riera, Raúl Pérez-Caballero, Silvina Natalini Martínez, Miriam González Amores, Laura Roig Soria, Laia Bernad, Santiago Cavero, Benjamin Trinité, Elena Aurrecoechea Pumar, Roc Pomarol Moyà, Borja Guarch-Ibáñez, Francesc Ripoll Oliveras, Silvia Narejos Perez, Cristina Calvo, Pyrene Martínez Piera, Julià Blanco, Montserrat Plana Prades, Julia G Prado, Pere Soler-Palacín, the RBDCOV study group

## Abstract

**Objectives:** Although rare, severe complications may follow SARS-CoV-2 infection in pediatric populations. PHH-1V, a protein-based, bivalent, adjuvanted COVID-19 vaccine, demonstrated strong immunogenicity and favorable safety as a booster in adults. This study further evaluated PHH-1V as a heterologous booster in pediatric adolescent population.

**Study design:** HIPRA-HH-3 was a phase IIb, open-label, multicenter, non-inferiority trial assessing the immunogenicity and safety of PHH-1V in 240 healthy adolescents (≥ 12 to < 18 years) previously vaccinated with BNT162b2. Immunogenicity (N = 88) was evaluated by measuring neutralizing antibodies against SARS-CoV-2 variants, total binding antibodies, and T-cell responses at day 14 post-booster, compared to young adults (≥ 18 to ≤ 25 years) (N = 81) from the previous HIPRA-HH-2 study, who had also received the PHH-1V booster vaccine. Safety endpoints included all adverse events through day 28.

**Results:** Neutralizing titers against Omicron BA.1 (primary endpoint), WH1, Beta, and Delta variants and total binding antibodies were significantly higher in adolescents than in young adults (p < 0.001), demonstrating non-inferiority (margin < 1.5). T-cell responses against Omicron BA.1 and Beta variants also increased significantly (p < 0.001). Adverse events were mostly mild or moderate, primarily injection-site pain and headache. No serious adverse events were reported.

**Conclusions:** PHH-1V is safe and immunogenic as a heterologous booster in adolescents, inducing both humoral and cellular responses with SARS-CoV-2 cross-variant immune recognition. These findings support the use of PHH-1V in adolescent vaccination strategies. Additional data on long-term protection in this population, including specific risk groups, remain needed.

**Clinical Trial Registration:** NCT06234956 (https://clinicaltrials.gov/study/NCT06234956) and 2023-504639-42-00 (https://euclinicaltrials.eu/search-for-clinical-trials/?lang=en&EUCT=2023-504639-42-00).

## Introduction

Since the onset of the COVID-19 pandemic, adult vaccination has markedly reduced the global disease burden by lowering the incidence of severe disease, particularly among high-risk populations[1,2]. Moreover, vaccination has played a crucial role in limiting viral transmission and severe clinical outcomes in younger populations[3,4].

Compared to adults, children and adolescents typically experience milder illness and more favorable outcomes, likely due to age-related differences in immune responses[5]. However, pediatric populations remain susceptible to severe pulmonary symptoms, long-term complications, and, in some cases, progression to multisystem inflammatory syndrome in children (MIS-C)[5–7]. Accordingly, recommendations for COVID-19 vaccination for healthy children and adolescents aged 6 months and older are based on shared clinical decision-making with healthcare providers, consistent with FDA-licensed indications in the United States (U.S.)[8]. In the EU, recommendations prioritize COVID-19 vaccination for children and adolescents with underlying conditions, with national authorities deciding on the inclusion of younger age groups[9].

Currently, up to three vaccines are authorized for children or adolescents in the U.S. and the EU[10,11]. Recommendations increasingly favor heterologous prime-boost strategies involving different vaccine platforms due to their ability to elicit stronger immune responses compared to homologous vaccination, while maintaining comparable safety profiles. Additionally, heterologous boosting strategies could potentially mitigate vaccine supply constraints and vaccine hesitancy[12,13]. While direct evidence in the pediatric population is still limited, findings from adult studies support the immunological advantages of heterologous booster approaches and suggest their potential applicability in children and adolescents[14].

PHH-1V is a recombinant protein-based vaccine comprising a fusion heterodimer of the receptor-binding domain (RBD) derived from the spike (S) protein of SARS-CoV-2 Beta and Alpha variants, adjuvanted with a squalene-based oil-in-water emulsion (SQBA)[15], with demonstrated safety and robust immunogenicity as a heterologous booster in adults[14]. PHH-1V COVID-19 vaccine received the recommendation for approval from the European Medicines Agency (EMA) for individuals aged 16 years and older[16]. This study aims to further evaluate the humoral and cellular immunogenicity, including cross-variant immune recognition of PHH-1V as a booster vaccine in adolescents aged ≥ 12 to < 18 years through immunobridging with young adults, and to assess its safety profile.

## Methods

### Study Design

HIPRA-HH-3 was a phase IIb, open-label, uncontrolled, single-agent, multicenter, non-inferiority clinical trial assessing the safety and immunogenicity of PHH-1V as a heterologous booster dose for COVID-19 prevention in adolescents aged ≥ 12 to < 18 years.

The protocol was approved on May 26, 2023 by the Independent Ethics Committee of HM Hospitales (CEIm HM Hospitales, Madrid, Spain) and the Spanish Agency of Medicines and Medical Devices (AEMPS). The study was conducted at seven sites across Spain (Supplementary Information, Section 2) in accordance with the 1975 Declaration of Helsinki, the International Council for Harmonization Good Clinical Practice guidelines, and applicable national regulations. Written informed consent was obtained from participants’ parents or legal guardians, and written assent was obtained from adolescents prior to enrollment. This study was posted on clinicaltrials.gov (NCT06234956) and at the [EU Clinical Trials Register](https://euclinicaltrials.eu/search-for-clinical-trials/?lang=en) (2023-504639-42-00). This manuscript adheres to the CONSORT reporting guidelines.

### Participants and Study Settings

Eligible participants were adolescents aged ≥ 12 to < 18 years at screening who had received two previous doses of the BNT162b2 vaccine at least six months before screening and tested negative for SARS-CoV-2 by rapid antigen test (RAT) at baseline (day 0). Key exclusion criteria included documented COVID-19 history (for the immunogenicity analyses), prior receipt of COVID-19 medications (except BNT162b2), receipt of non-study vaccines within 14 days before or after screening, and any live or attenuated SARS-CoV-2 vaccine within 4 weeks before or after screening. Complete eligibility criteria and data collection methods are detailed in Supplementary Information, Sections 3 and 4, respectively.

The total clinical follow-up period per participant was 12 months. The HIPRA-HH-3 trial was initiated on June 8, 2023 (first participant, first visit) and was completed on September 9, 2025 (last participant, last visit). This manuscript presents data through day 28 post-booster.

### Interventions

The PHH-1V vaccine was supplied in ready-to-use, 0.5 mL single-dose vials stored at 2–8 °C, containing 40 μg of recombinant protein (selvacovatein, active substance) adjuvanted with SQBA, produced by HIPRA SCIENTIFIC, S.L.U. (Amer, Spain). At day 0, participants received an intramuscular injection in the non-dominant arm. The selected dose and schedule were approved by EMA’s Pediatric Committee (P/0465/2022) (Supplementary Information, Section 4).

### Outcomes

The primary efficacy endpoint was the neutralizing antibody titer against Omicron BA.1, measured 14 days after administering the PHH-1V booster vaccine in adolescents, compared to young adults (aged ≥ 18 to ≤ 25 years) from the previous HIPRA-HH-2 study, in which enrollment began in November 2021 and follow-up concluded in May 2023[14]. Secondary efficacy endpoints reported here were the neutralizing titers against other SARS-CoV-2 variants (original WH1 [WH1], B.1.351 [Beta], B.1617.2 [Delta]), total binding antibody titers against SARS-CoV-2 RBD, geometric mean fold-rise (GMFR) in neutralizing and total binding antibodies, percentage of participants with ≥ 4-fold increase in binding antibody titers, and RBD-specific T-cell responses against Omicron BA.1 and Beta variants in a subset of approximately 10% of participants. This manuscript presents endpoint results from days 0 to 14 post-booster, including complementary analyses described in Supplementary Information, Section 5.

Neutralizing antibody responses were measured by pseudovirion-based neutralization assay (PBNA). Total RBD-binding antibodies were analyzed by electrochemiluminescence immunoassay (ECLIA). Cellular immunogenicity assays included interferon-gamma (IFN-γ) enzyme-linked immunosorbent spot (ELISpot) and intracellular cytokine staining (ICS) for CD4^+^ and CD8^+^ T cells using participant-derived cryopreserved peripheral blood mononuclear cells (PBMCs) isolated by ficoll-density gradient from 10 mL of whole blood samples. The PBNA[17,18], ECLIA[14], and cellular immunogenicity assays[14] were performed as previously described (Supplementary Information, Section 5). Additionally, *CXCL10* gene expression from 1 mL of fresh whole blood was quantified by RT-qPCR as an indirect readout of IFN-γ^+^ T-cell reactivity against SARS-CoV-2 viral antigens (Supplementary Information, Section 5).

Primary safety endpoints reported here included treatment-emergent adverse events (TEAEs), i.e., solicited local and systemic reactions through day 7 and unsolicited adverse events (AEs) through day 28 (Supplementary Information, Section 4).

### Sample Size and Statistical Methods

We compared the antibody geometric mean titer (GMT) in adolescents with that in young adults from the previous HIPRA-HH-2 study[14], using a 1.5-fold non-inferiority margin for immunobridging analysis. With 88 adolescents assessed, the study provided 81.5% statistical power, surpassing the recommended 80% threshold[19].

Immunogenicity was measured using GMT and GMFR (day 14/day 0 antibody titers), analyzed through mixed-effects models and Student’s t-tests. Non-inferiority was based on 95% confidence intervals (CIs) for GMT ratios[20]. Cellular immunity responses were analyzed by paired t-tests and mixed-effects models. Statistical significance was set at p < 0.05. Statistical analyses and visualizations were conducted using SAS (version 9.4), R (version 4.1.3 or later), and GraphPad Prism (version 10.4.2) (Supplementary Information, Section 6).

## Results

### Study Population

Of 242 enrolled participants, two were excluded at screening due to not meeting all inclusion criteria. The intention-to-treat (ITT) population and safety population (SP) each consisted of 240 participants who received the PHH-1V booster vaccine. One additional participant was later excluded from the ITT due to not meeting all inclusion criteria, thus defining the modified intention-to-treat (mITT) population. Eighty-eight participants from the mITT were included in the immunogenicity population (IGP) to evaluate the humoral response against Omicron BA.1, WH1, Beta, and Delta variants through day 14 post-booster. A subanalysis of the humoral response against Omicron XBB.1.5 and XBB.1.16 was performed using PBNA with available samples from 44 participants. T-cell response was analyzed by ELISpot and ICS in samples from 16 adolescents (Fig 1). Participant baseline characteristics for SP, IGP, and the young adult control group are presented in Table 1. Baseline characteristics of study participants included in the cellular immunogenicity analysis against Omicron BA.1 and Beta variants (ELISpot and ICS) and in the XBB variant subanalysis (PBNA) are shown in Supplementary Table 1.

**Figure 1:**
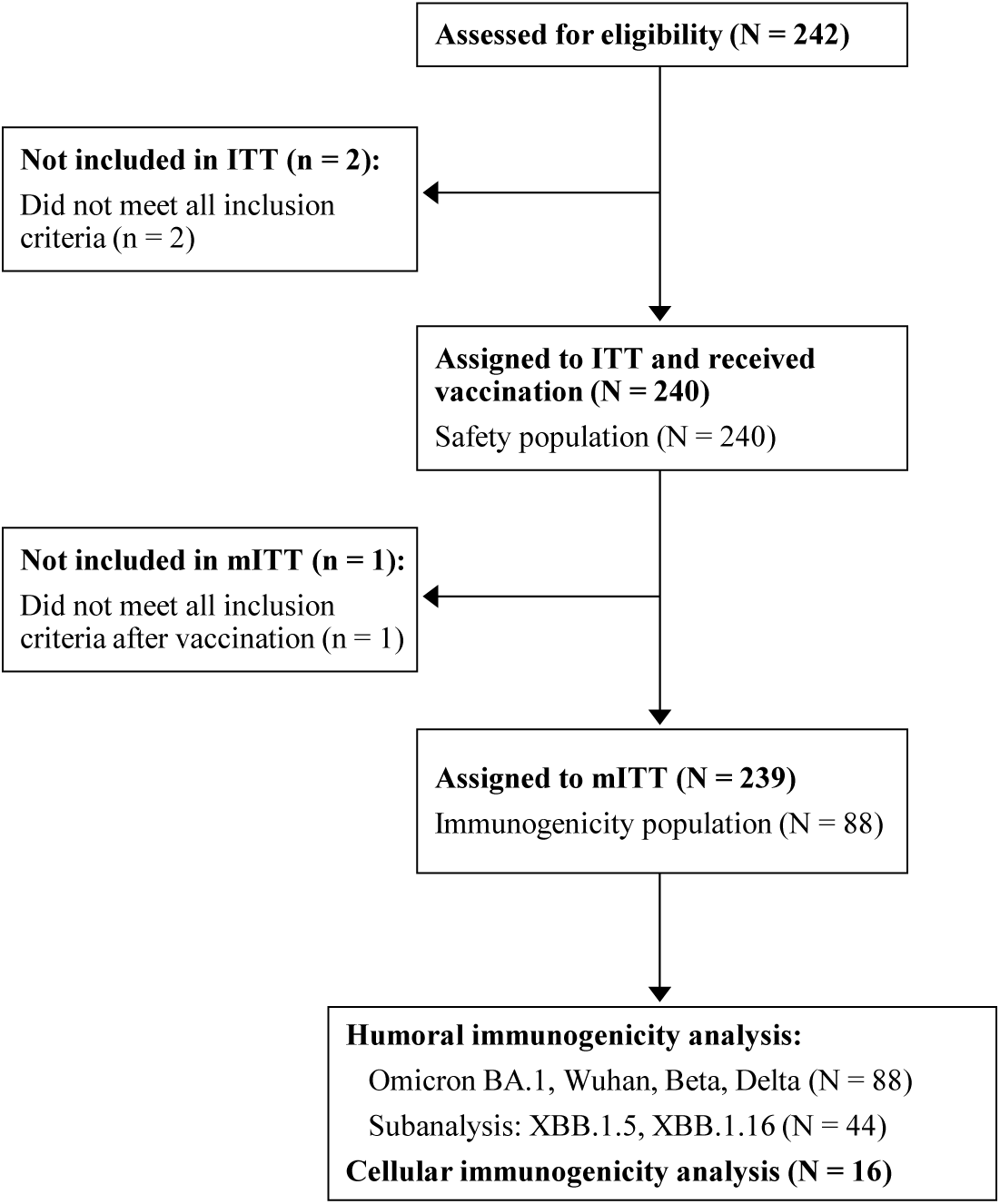
Study flowchart and participant disposition.

**Table 1.**
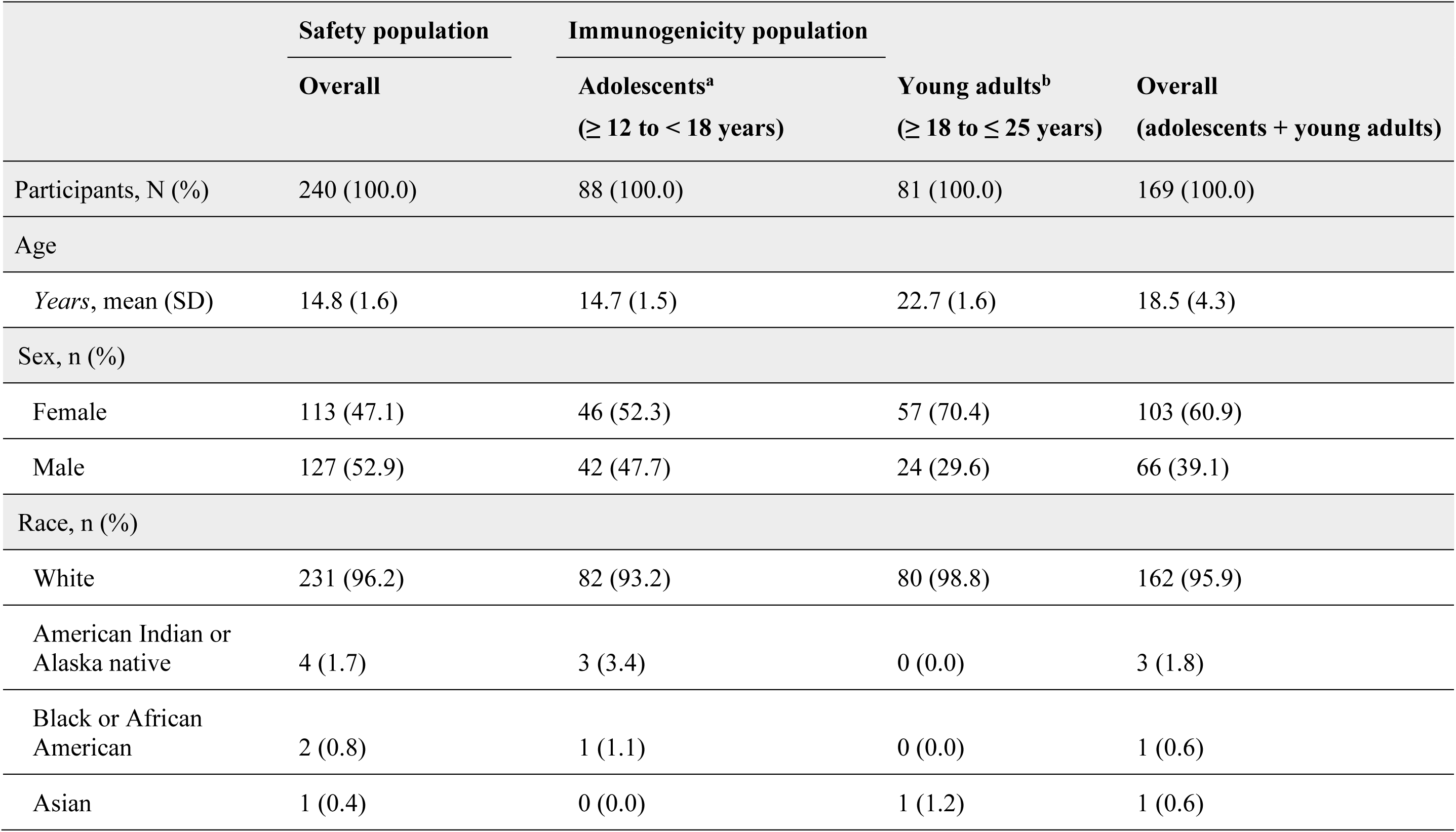
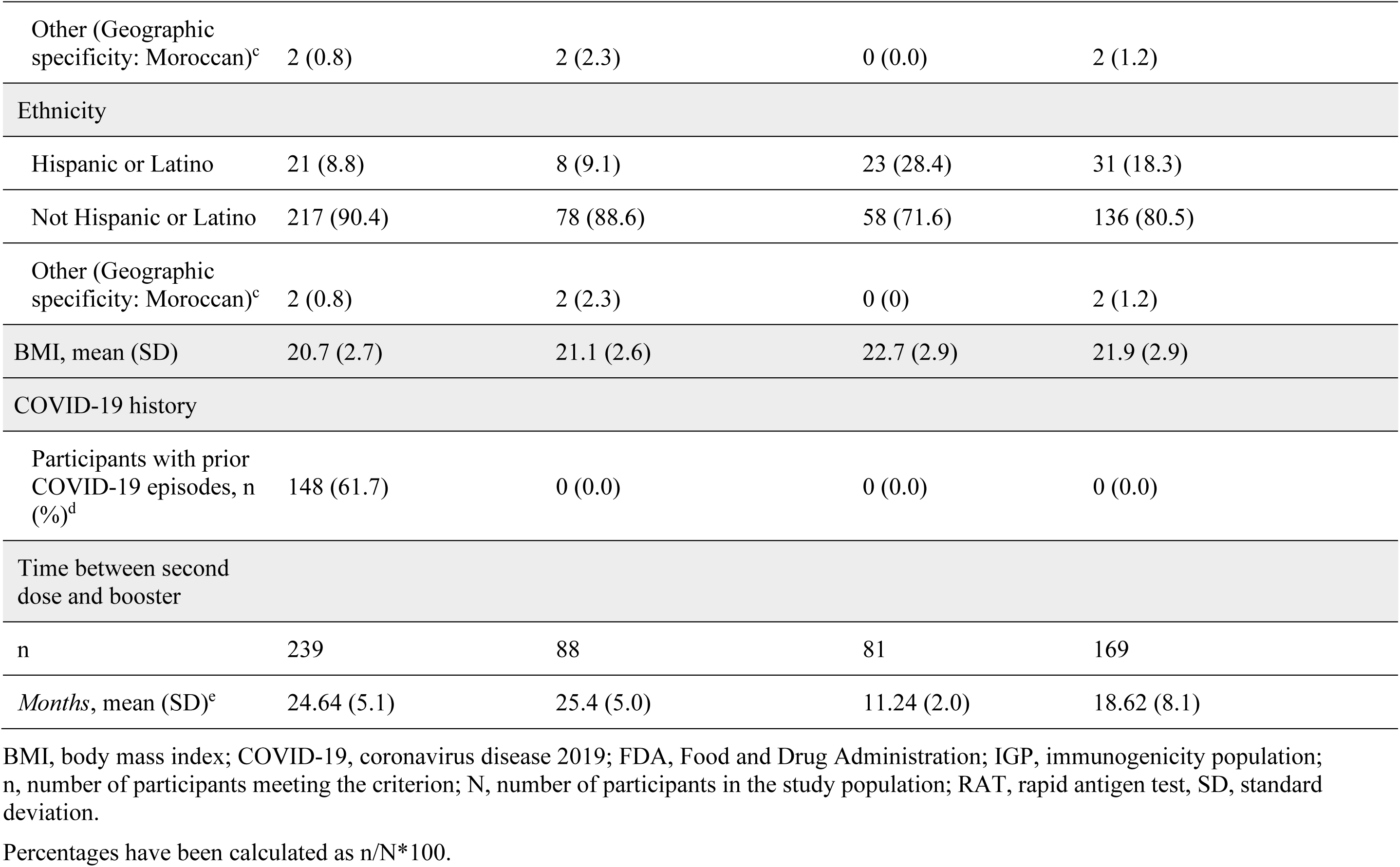
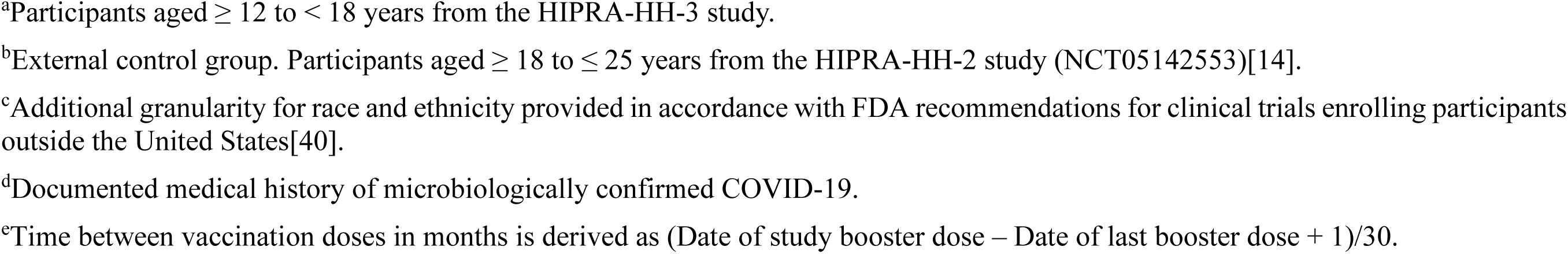
Baseline characteristics of study participants.

|  | Safety population | Immunogenicity population |  |  |
| --- | --- | --- | --- | --- |
|  | Overall | Adolescents<sup>a</sup><br>(≥ 12 to < 18 years) | Young adults<sup>b</sup><br>(≥ 18 to ≤ 25 years) | Overall<br>(adolescents + young adults) |
| Participants, N (%) | 240 (100.0) | 88 (100.0) | 81 (100.0) | 169 (100.0) |
| Age |  |  |  |  |
| *Years*, mean (SD) | 14.8 (1.6) | 14.7 (1.5) | 22.7 (1.6) | 18.5 (4.3) |
| Sex, n (%) |  |  |  |  |
| Female | 113 (47.1) | 46 (52.3) | 57 (70.4) | 103 (60.9) |
| Male | 127 (52.9) | 42 (47.7) | 24 (29.6) | 66 (39.1) |
| Race, n (%) |  |  |  |  |
| White | 231 (96.2) | 82 (93.2) | 80 (98.8) | 162 (95.9) |
| American Indian or<br>Alaska native | 4 (1.7) | 3 (3.4) | 0 (0.0) | 3 (1.8) |
| Black or African<br>American | 2 (0.8) | 1 (1.1) | 0 (0.0) | 1 (0.6) |
| Asian | 1 (0.4) | 0 (0.0) | 1 (1.2) | 1 (0.6) |
| Other (Geographic specificity: Moroccan)<sup>c</sup> | 2 (0.8) | 2 (2.3) | 0 (0.0) | 2 (1.2) |
| Ethnicity |  |  |  |  |
| Hispanic or Latino | 21 (8.8) | 8 (9.1) | 23 (28.4) | 31 (18.3) |
| Not Hispanic or Latino | 217 (90.4) | 78 (88.6) | 58 (71.6) | 136 (80.5) |
| Other (Geographic specificity: Moroccan)<sup>c</sup> | 2 (0.8) | 2 (2.3) | 0 (0) | 2 (1.2) |
| BMI, mean (SD) | 20.7 (2.7) | 21.1 (2.6) | 22.7 (2.9) | 21.9 (2.9) |
| COVID-19 history |  |  |  |  |
| Participants with prior COVID-19 episodes, n (%)<sup>d</sup> | 148 (61.7) | 0 (0.0) | 0 (0.0) | 0 (0.0) |
| Time between second dose and booster |  |  |  |  |
| n | 239 | 88 | 81 | 169 |
| *Months*, mean (SD)<sup>c</sup> | 24.64 (5.1) | 25.4 (5.0) | 11.24 (2.0) | 18.62 (8.1) |

BMI, body mass index; COVID-19, coronavirus disease 2019; FDA, Food and Drug Administration; IGP, immunogenicity population; n, number of participants meeting the criterion; N, number of participants in the study population; RAT, rapid antigen test, SD, standard deviation.
Percentages have been calculated as $n/N \times 100$ .
<sup>a</sup>Participants aged $\geq 12$ to $< 18$ years from the HIPRA-HH-3 study.
<sup>b</sup>External control group. Participants aged $\geq 18$ to $\leq 25$ years from the HIPRA-HH-2 study (NCT05142553)[14].
<sup>c</sup>Additional granularity for race and ethnicity provided in accordance with FDA recommendations for clinical trials enrolling participants outside the United States[40].
<sup>d</sup>Documented medical history of microbiologically confirmed COVID-19.
<sup>e</sup>Time between vaccination doses in months is derived as (Date of study booster dose – Date of last booster dose + 1)/30.

### Humoral Responses to SARS-CoV-2 Variants After PHH-1V Booster

At day 0, adjusted neutralizing antibody GMT [95% CI] against Omicron BA.1 in adolescents and young adults was 1303.54 [1016.05; 1672.39] and 48.48 [37.39; 62.86], respectively. By day 14, adjusted GMT [95% CI] increased to 24081.34 [19741.36; 29375.43] in adolescents and 2340.57 [1900.42; 2882.66] in young adults (Fig 2A and Supplementary Table 2). The GMT ratio (young adults vs. adolescents) at day 14 was 0.10 [0.07; 0.13] (p < 0.001), indicating not only non-inferiority but a superior neutralizing antibody titer against Omicron BA.1 in adolescents compared to young adults (Fig 2B and Supplementary Table 2) as primary efficacy endpoint.

**Figure 2:**
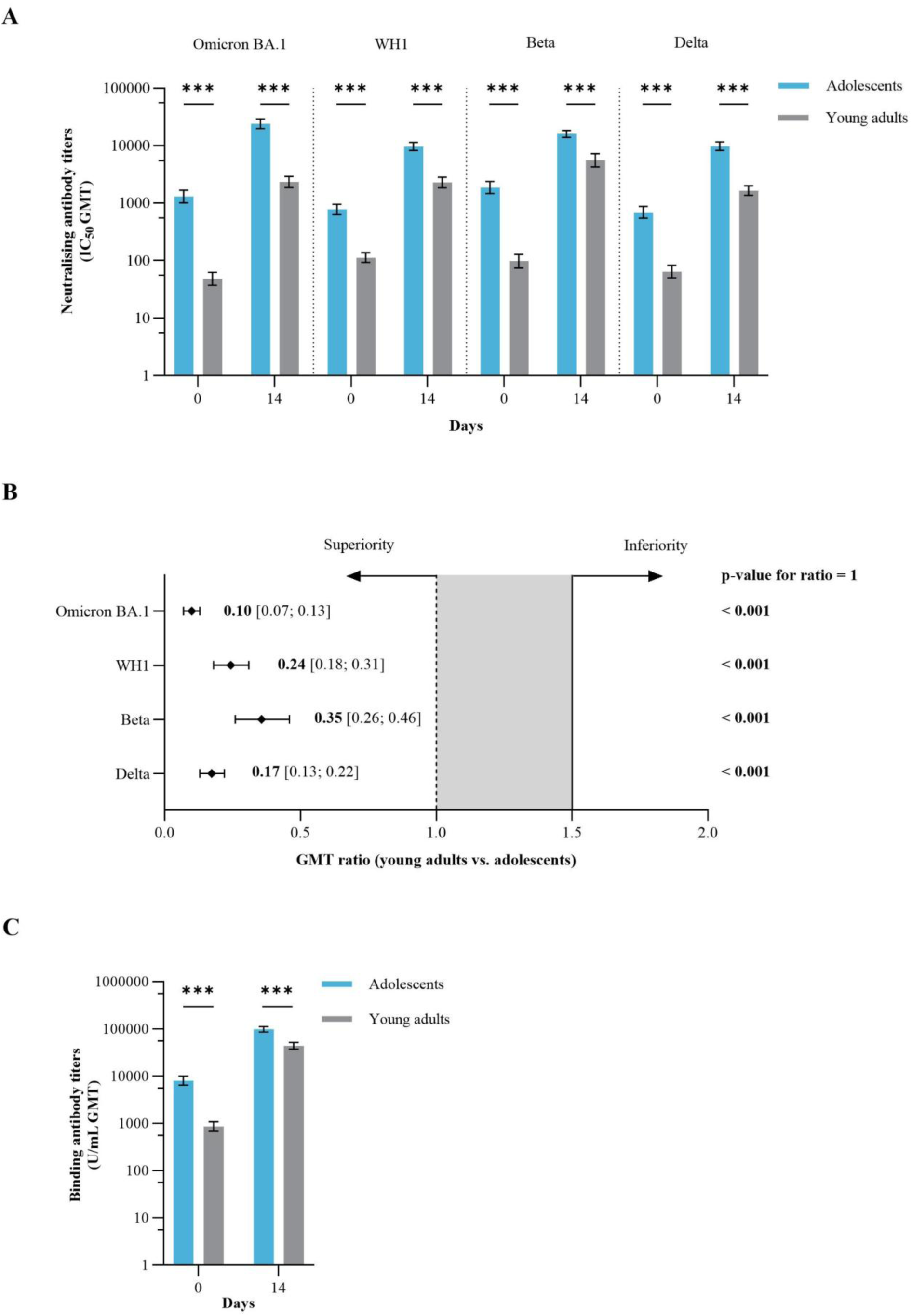
Humoral immunogenicity in response to PHH-1V booster vaccine. **(A)** Neutralizing antibody titers against SARS-CoV-2 variants in adolescents (N = 88) vs. young adults (N = 81) at day 0 and day 14 post-booster. **(B)** Forest plot for neutralizing antibody GMT ratio (95% CI) in young adults vs. adolescents at day 14. The solid line represents the non-inferiority margin (1.5), and the dashed line indicates the superiority limit (GMT ratio = 1). **(C)** SARS-CoV-2 binding antibody titers in adolescents (N = 87, as one sample volume was insufficient for the analysis) vs. young adults (N = 81) at day 0 and day 14. Young adults (aged ≥ 18 to ≤ 25 years; HIPRA-HH-2 study [Corominas *et al.*[14]]) served as the external control group. Plots show GMTs with 95% CIs. Statistical analyses employed a mixed-effects model for repeated measures. ***p < 0.001.

As secondary efficacy endpoints, we tested cross-variant neutralizing and total antibody responses induced by PHH-1V booster. At day 14 post-vaccination, adolescents showed increased neutralizing titers against WH1, Beta, and Delta variants (Supplementary Fig A and Supplementary Table 2), and against XBB.1.5 and XBB.1.16 variants (Supplementary Fig B and Supplementary Table 2) compared to pre-booster levels, as measured by GMFR. Total RBD-binding antibody titers in adolescents also increased at day 14, with an associated GMFR [95% CI] of 12.39 [9.80; 15.67] (Supplementary Fig C and Supplementary Table 2). Overall, 73 (83.9%, 95% CI: [74.5; 90.9]) adolescent participants achieved a ≥ 4-fold increase in binding antibody titers post-vaccination boost.

Additional analyses compared the humoral responses against WH1, Beta, Delta, and Omicron XBB variants between adolescents and young adults. At day 14, adolescents showed significantly higher neutralizing antibody GMTs against WH1, Beta, and Delta compared to young adults (p < 0.001) (Fig 2A and Supplementary Table 2). The values of neutralizing GMT ratios demonstrated non-inferiority in adolescents compared to young adults for all tested variants (p < 0.001) (Fig 2B and Supplementary Table 2). In contrast, GMFR values for neutralizing antibodies against Omicron BA.1, WH1, Beta, and Delta variants were significantly lower in adolescents than in young adults (p < 0.001, p < 0.01, p < 0.001, and p < 0.01, respectively) (Supplementary Fig A and Supplementary Table 2).

At days 0 and 14, adolescents showed significantly higher total binding antibody GMTs compared to young adults (p < 0.001) (Fig 2C and Supplementary Table 2). However, GMFR values for total binding antibodies were significantly lower in adolescents than in young adults (p < 0.001) (Supplementary Fig C and Supplementary Table 2).

### Cellular Responses to Omicron BA.1 and Beta SARS-CoV-2 Variants After PHH-1V Booster

In terms of cellular responses, adolescents receiving PHH-1V showed a significant increase in IFN-γ-producing lymphocytes in response to Omicron BA.1 and Beta variants peptide pools by day 14 (p < 0.001) (Fig 3A). All participants were T-cell responders to the vaccination against both variants (16 responders; 100%; 95% CI: [79.4; 100]).

**Figure 3:**
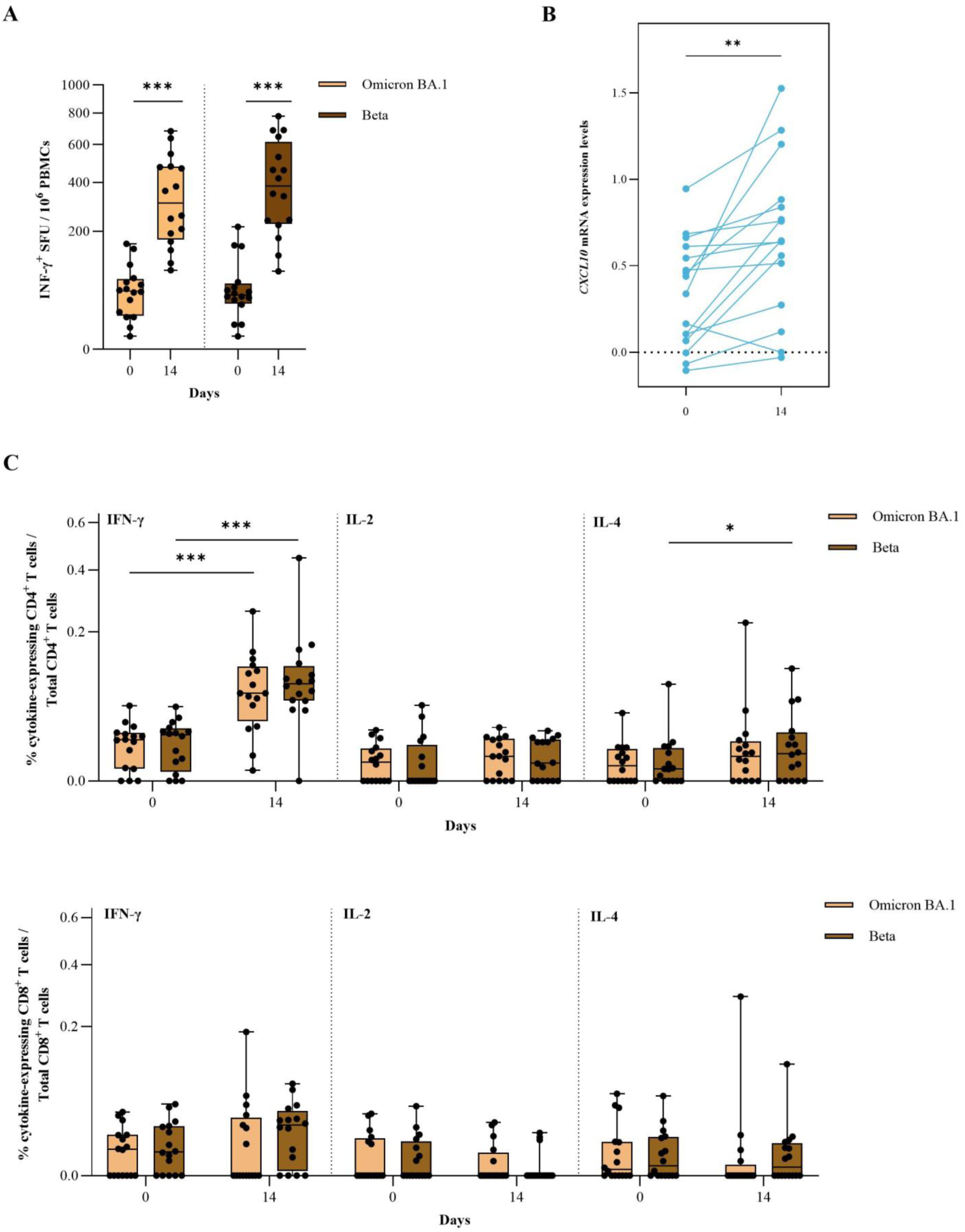
Cellular immunogenicity in response to PHH-1V booster vaccine. T-cell responses were assessed in adolescents at days 0 and 14 post-booster (N = 16) using IFN-γ^+^ ELISpot **(A)**, *CXCL10* quantification by RT-qPCR **(B)**, and CD4^+^/CD8^+^ ICS of lymphocytes secreting IFN-γ, IL-2, and IL-4 **(C)**. For ELISpot and ICS, PBMCs were stimulated with RBD peptide pools from Omicron BA.1 and Beta SARS-CoV-2 variants. *CXCL10* mRNA levels were used as a surrogate marker of IFN-γ-mediated T-cell activity in whole blood samples stimulated with a peptide pool covering SARS-CoV-2 S protein (N = 16). Boxes show the IQR, with medians depicted as solid lines and whiskers extending 1.5 x IQR. IQR, interquartile range. Statistical analyses were conducted using paired-samples t-tests. *p < 0.05; **p < 0.01; ***p < 0.001.

We additionally evaluated the potential of measuring *CXCL10* mRNA expression in whole blood samples from adolescents as an indirect readout of T-cell immunity. We found a significant increase between day 0 and day 14 that further confirmed PHH-1V-induced T-cell activation in this population (p = 0.002) (Fig 3B).

To complete the functional characterization of CD4^+^ and CD8^+^ T-cell responses after PHH-1V booster vaccination, we performed ICS of PBMCs. Our results revealed a significant increase in IFN-γ-expressing CD4^+^ T cells at day 14 following stimulation with RBD derived peptide pools from Omicron BA.1 (p < 0.001) and Beta (p < 0.001) variants, as well as an increase in interleukin-4 (IL-4)-producing CD4^+^ T cells against Beta (p = 0.04) (Fig 3C). No significant changes were observed in interleukin-2 (IL-2)-producing CD4^+^ or in CD8^+^ T cells for any of the variants tested (Fig 3C).

### Safety and Tolerability

A total of 206 (85.8%) participants reported at least one TEAE, most of which were mild or moderate. Of these, 198 (82.5%) participants reported vaccine-related TEAEs. The most common TEAEs were injection site pain (187 participants; 77.9%) and headache (77; 32.1%) (Fig 4 and Supplementary Table 3). No serious adverse events, medically attended adverse events, adverse events of special interest, or deaths were reported during the 28-day post-booster period covered by this interim study analysis.

**Figure 4:**
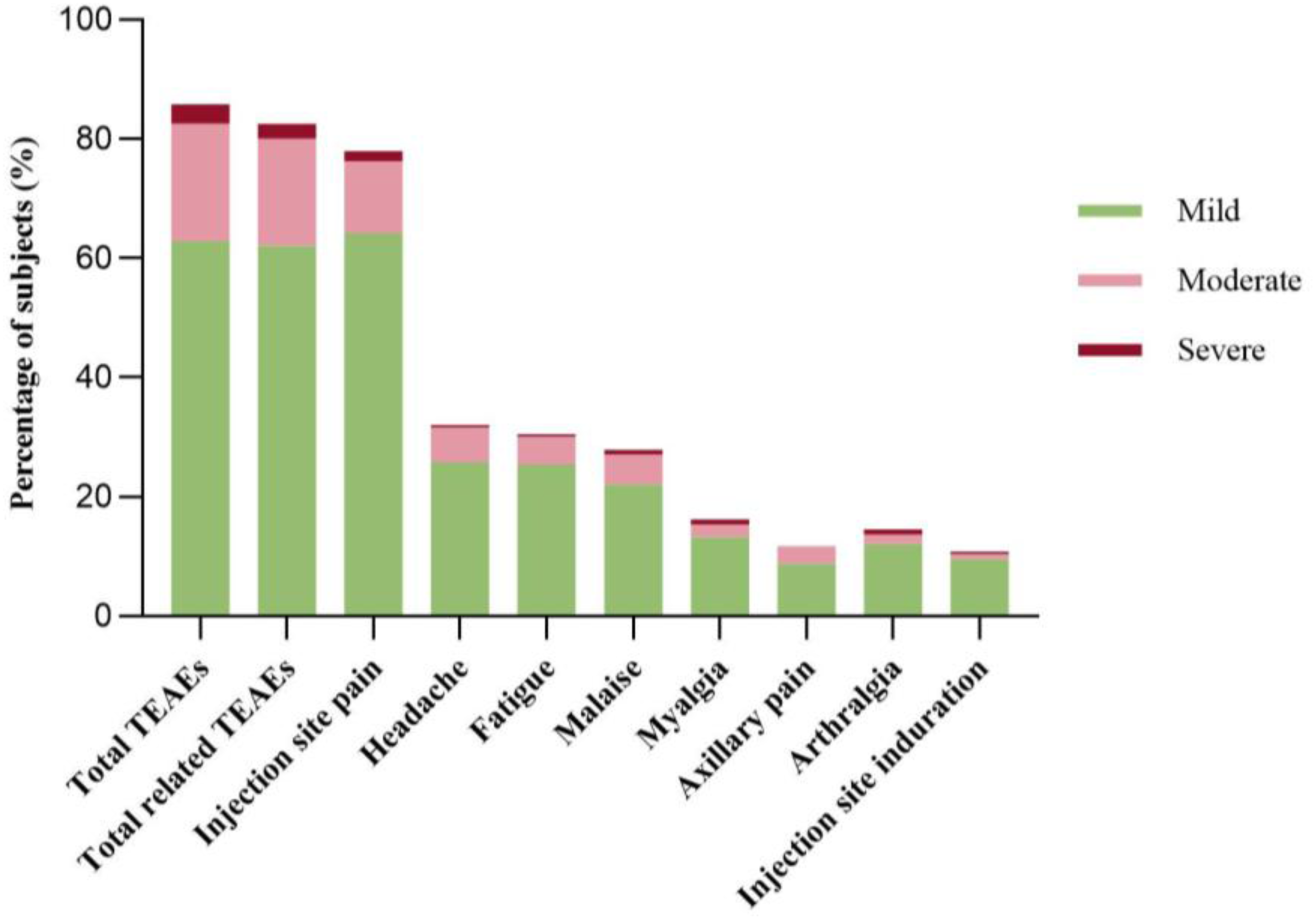
Frequency and severity of TEAEs through day 28 post-booster. TEAEs by MedDRA PT reported in ≥ 10.0% of participants through day 28 post-booster. Percentages are relative to SP (N = 240), indicating TEAE intensity (mild, moderate, and severe). Participants with multiple TEAEs are counted once per PT as the most severe event. MedDRA, Medical Dictionary for Regulatory Activities; PT, preferred term.

The most frequent solicited local reactions through day 7 were pain at the injection site (174 participants; 72.5%) and tenderness (134; 55.8%). Headache (77; 32.1%), fatigue (73; 30.4%), and malaise (70; 29.2%) were the most common solicited systemic events, which gradually declined through day 7. The solicited and remaining unsolicited events through day 28 are shown in Supplementary Table 4-6.

## Discussion

The emergence of new SARS-CoV-2 variants highlights the need for heterologous vaccination booster approaches, particularly in pediatric populations, where clinical data remain limited. The HIPRA-HH-3 study addresses this evidence gap by evaluating PHH-1V’s immunogenicity and safety as a heterologous booster in adolescents, building on prior information from PHH-1V’s adult trials.

For the present study, we leveraged prior evidence from phase IIb[14] and phase III[21] trials conducted in participants aged ≥ 18 and ≥ 16 years, respectively, to inform our selection of vaccine dose and schedule. Specifically, we adopted the adult-approved vaccine dose (40 μg of selvacovatein) and a 6-month boosting interval[16], in alignment with regulatory considerations at the time of study initiation[22].

The study met its primary endpoint, with adolescents showing neutralizing titers against Omicron BA.1 that not only met non-inferiority criteria but also exceeded those reported in young adults from the previous HIPRA-HH-2 study[14]. Interestingly, the corresponding GMFR for both neutralizing antibodies across all tested variants and total binding antibodies was lower in adolescents than in young adults, probably due to the lower baseline levels observed in the latter group. Several factors may account for this observation. First, enhanced (1.76-fold) humoral responses to two doses of BNT162b2 vaccine have been reported in adolescents (aged 12-15 years) compared to young adults (aged 16-25 years)[23]. This observation, combined with the slower decay of antibodies in adolescents compared to young individuals[24], may have contributed to the observed baseline differences. However, epidemiological differences between the recruitment periods across study groups (HIPRA-HH-3 adolescents and HIPRA-HH-4 young adults) may explain the observed differences. To ensure full comparability, both groups were recruited using similar inclusion criteria (no recorded SARS-CoV-2 infection and two previous doses of mRNA vaccines); however, vaccination started in November 2021 for young adults and in June 2023 for adolescents. This interval encompasses the Omicron wave, with global seropositivity increasing from 16% in February 2021 to 67% by October 2021[25]. Epidemiological data from the Catalonia region (Spain) indicated seroprevalence rates of 98-99% among adolescents as of May 2022[26]. Hence, a widespread subclinical exposure to SARS-CoV-2 Omicron subvariants might explain the enhanced baseline immunity levels in adolescents.

Importantly, our study demonstrates broad neutralizing activity induced by PHH-1V, a SARS-CoV-2 Beta-Alpha heterodimer, against multiple variants of concern. The highly transmissible Omicron BA.1 variant—selected to define the primary endpoint— was rapidly overtaken by newer Omicron sublineages, including the highly immune-evasive XBB recombinant lineage[27,28]. By early 2023, the XBB.1.5 subvariant rose to predominance, accounting for nearly 60% of cases in the U.S. by May 2023, subsequently spreading internationally[29,30]. Throughout late 2023 and into 2024, XBB descendants remained dominant[31]. The observed cross-variant neutralization against all tested variants, including the XBB lineage, strongly indicates the potential of PHH-1V to confer anticipatory cross-protection against newly emerging SARS-CoV-2 strains. Notably, the PHH-1V vaccine design enables rapid antigenic adaptation, effectively addressing evolving SARS-CoV-2 variants. This is demonstrated by the subsequent development and EU approval of the monovalent Omicron XBB.1.16-adapted PHH-1V81 vaccine[16,32], following regulatory recommendations to update vaccine compositions with XBB-lineage variants[33], which were published after the approval of this study protocol.

Furthermore, our findings indicate that PHH-1V, administered as a heterologous booster, effectively induced SARS-CoV-2 T-cell responses—mainly of the IFN-γ^+^ CD4^+^ phenotype— at two weeks post-booster following exposure to the Omicron BA.1 and Beta variants. These findings are in agreement with previous studies in adults[14]. Importantly, the vaccine’s capacity to stimulate both humoral and cellular arms of adaptive immunity contributes to cross-variant protection and long-lasting immune memory[34,35], supporting COVID-19 vaccine strategies in the pediatric population. Additionally, our findings, consistent with recent work utilizing *CXCL10* gene expression as an indirect readout of T-cell immunity in children[36], highlight a promising immune monitoring tool requiring small-volume samples. This is particularly valuable for pediatric studies involving younger children, for whom obtaining larger blood volumes is challenging.

Moreover, the favorable safety and reactogenicity profile observed for PHH-1V in adolescents further supports its potential inclusion in pediatric vaccination strategies. Most reported TEAEs were mild or moderate, typically limited to injection-site pain and headache, without any serious events. This aligns well with the reactogenicity profiles observed in adults, which were generally mild-to-moderate, including local injection-site pain, headache, and fatigue[14]. Importantly, some systemic AEs frequently observed in adolescents receiving one or two doses of other COVID-19 vaccines[37,38] were reported at low rates among adolescent participants receiving PHH-1V. Arthralgia and pyrexia were reported in 14.6% and 3.8% of participants, respectively. Chills were not reported. Hence, PHH-1V’s favorable reactogenicity profile may help to reduce vaccine hesitancy and encourage acceptance among adolescents and their families.

Nonetheless, this study has some limitations. Although participants with recorded prior SARS-CoV-2 infection were actively excluded following the criteria defined in the HIPRA-HH-2 trial[14], the absence of complete baseline serological screening (anti-S and anti-nucleocapsid (N) protein antibodies) could not definitively rule out prior asymptomatic infections, introducing potential confounding factors into immunogenicity analyses. However, the fast decay of anti-N antibodies, which typically become undetectable in adolescents just a few weeks after infection[39], would also prevent reliable serological detection of past infections. Additionally, no long-term protection data are presented in this interim study analysis.

Ongoing follow-up within the HIPRA-HH-3 study over one year will provide critical insights into the sustained immunogenicity and safety of the vaccine, supporting informed recommendations for adolescent booster vaccinations in the context of evolving SARS-CoV-2 epidemiological dynamics.

In conclusion, PHH-1V induced a robust neutralizing antibody response against Omicron BA.1 and all other SARS-CoV-2 variants assessed in this study, ranging from the ancestral WH1 strain to the highly mutated and distant Omicron XBB variants. It also significantly boosted SARS-CoV-2-specific T-cell responses and demonstrated a favorable safety and tolerability profile when administered as a heterologous booster following two doses of BNT162b2 in adolescents. These findings underscore the potential of PHH-1V as a beneficial addition to existing authorized COVID-19 booster options, particularly considering the continuous emergence of new viral variants and the ongoing need for broadened immune protection.

## CRediT authorship contribution statement

**Ignasi Esteban Riera; Montserrat Plana Prades; Elena Aurrecoechea Pumar; Roc Pomarol Moyà; Laia Bernad; Santiago Cavero; Raúl Pérez-Caballero; Julia G. Prado:** Investigation; Formal analysis; Writing – review & editing. **Laura Roig Soria:** Investigation; Formal analysis; Data curation; Writing – review & editing. **Benjamin Trinité; Julià Blanco; Silvina Natalini Martínez; Cristina Calvo; Borja Guarch-Ibáñez; Francesc Ripoll Oliveras; Pyrene Martínez Piera; Silvia Narejos Perez:** Investigation; Writing – review & editing. **Pere Soler-Palacín; Miriam González Amores:** Conceptualization; Methodology; Project administration; Supervision; Data curation; Writing – original draft; Writing – review & editing. All authors critically revised the manuscript for important intellectual content, approved the final version, and agree to be accountable for all aspects of the work.

## Acknowledgements

We thank Adriana Margarit Soler, Antoni Soriano Arandes, Beatriz Álvarez Vallejo, Noemí Giménez Sanz, and Stephany Zelada for their active participation in the recruitment and follow-up of study participants (Hospital Universitari Vall d’Hebron); Alexandra Moros (HIPRA) for her contribution to managing and coordinating the procedures required to evaluate T-cell activity through *CXCL10* quantification by RT-qPCR; Anna Moya and Ruth Peña (HIPRA) for their technical assistance with the laboratory work; Anna Granés (HIPRA) for her support with data analysis and the preparation of figures and tables; Neus Cantariño (HIPRA) for her support with scientific writing; and the European Commission for its funding support.

## Declaration of Generative AI and AI-assisted technologies in the writing process

During the preparation of this work the authors used AI generative tools (ChatGPT^TM^, OpenAI, San Francisco, CA, USA) in order to improve language and readability of the text and as an aid to perform bibliographic searches. After using this tool, the authors reviewed and edited the contentas needed and take full responsibility for the content of the publication.

## Prior presentation of study data

Information from the HIPRA-HH-3 study has been previously presented at scientific congresses:

- Gil de Miguel, A. Características de la vacuna Bimervax [Internet]. Asociación Española de Vacunología (AEV) Symposium 2023; 2023 Nov 10; Palma de Mallorca, Spain.
- Natalini S, et al. Safety and immunogenicity study of PHH-1V adjuvanted protein vaccine used as heterologous booster in adolescents from 12 to 18 years of age. ePoster 2036. 42nd Annual Meeting of the European Society for Paediatric Infectious Diseases (ESPID); 2024 May 20; Copenhagen, Denmark.
- Esteban I, et al. Evaluation of the PHH-1V COVID-19 Vaccine Induced T-cell Responses in Adolescents: HIPRA-HH-3 Study. 3nd Conference on Retroviruses and Opportunistic Infections (CROI); 2025 Mar 9; San Francisco, US.

## Data sharing statement

Deidentified individual participant data (including data dictionaries), along with the study protocol, statistical analysis plan, and informed consent form, will be made available upon publication. Access will be granted to researchers who submit a methodologically sound proposal, subject to approval by HIPRA. Proposals should be submitted to

## Abbreviations

AE: adverse event
AESIs: adverse events of special interest
CI: confidence interval
ELISpot: enzyme-linked immunosorbent spot
GMFR: geometric mean fold rise
GMT: geometric mean titer
ICS: intracellular cytokine staining
IFN-γ: interferon-gamma
IGP: immunogenicity population
IL-2: interleukin-2
IL-4: interleukin-4
MMRM: mixed-effects model for repeated measures
PBMCs: peripheral blood mononuclear cells
PBNA: pseudovirion-based neutralization assay
RBD: receptor-binding domain
SAEs: serious adverse events
SD: standard deviation
SFU: spot-forming units
TEAEs: treatment-emergent adverse events

