## Supplementary figures and images for "Immunogenicity and Safety of PHH-1V COVID-19 Vaccine as a Heterologous Booster in Adolescents"

### Supplementary Figure 1

**Supplementary Figure 1**

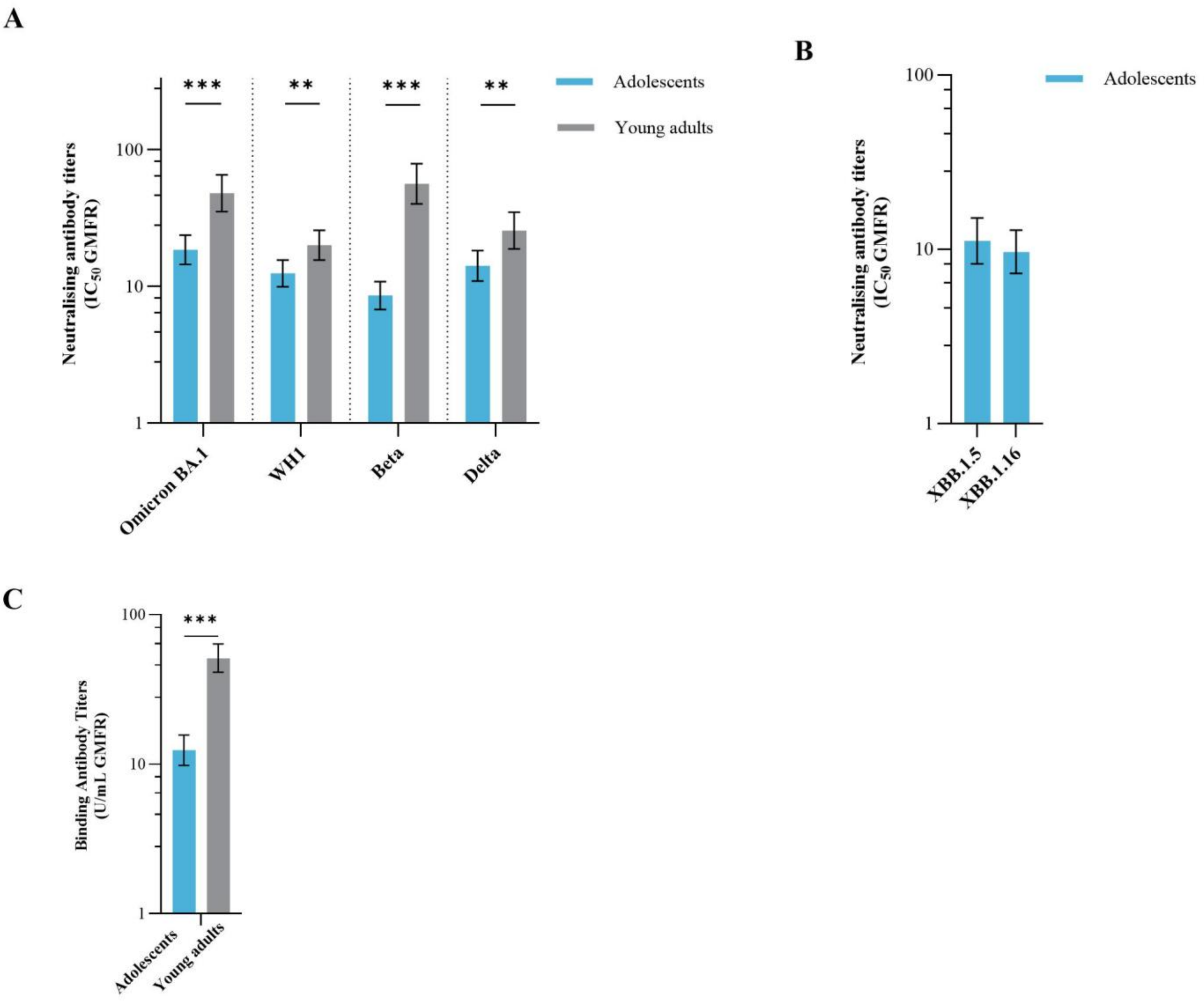
