## Supplementary Material for "Immunogenicity and Safety of PHH-1V COVID-19 Vaccine as a Heterologous Booster in Adolescents"

**Supplementary Information**

### 1. Members of the RBDCOV Group

HIPRA
Avinguda de la Selva, 135
17170 Amer, Girona, Spain

Members:
Alexandra Moros Sanz
Anna Granés Coll
Javiera González
Jon Insunza Gómez
Júlia Corominas García
Irina Güell Casellas
Laura Ferrer Soler
Lluís Riera Sans
Maria Glòria Pujol Prat
Maria Teresa Prat Cabañas
Meritxell Deulofeu Figueras
Neus Cantariño Iglesias
Toni Prenafeta Amargós

Hospital Universitari Vall d'Hebron
Passeig de la Vall d'Hebron, 119,
08035 Barcelona, Catalonia, Spain

Members:
Adriana Margarit Soler
Antoni Soriano Arandes
Beatriz Álvarez Vallejo
Noemí Giménez Sanz
Stephany Zelada

IrsiCaixa
Passeig de la Vall d'Hebron, 119,
08035 Badalona, Catalonia, Spain

Ruth Peña Ponderos
Julià Blanco

### 2. List of Participating Study Centers

Hospital Universitari Vall d'Hebron

Passeig de la Vall d'Hebron, 119,
08035 Barcelona, Catalonia, Spain

Principal Investigator:
Dr. Pere Soler Palacín

(+34) 934 89 31 00 (ext. 3378)

Hospital Universitari Dr. Josep Trueta

Avinguda de França, s/n,
17007 Girona, Catalonia, Spain

Principal Investigator:
Dr. Borja Guarch Ibáñez

(+34) 972 94 02 00 (ext 2574)

Hospital HM Montepríncipe

Av. de Montepríncipe, 25,
28660 Boadilla del Monte, Madrid, Spain

Principal Investigator:
Dr. Silvina Natalini Martínez


Hospital la Paz

Paseo de la Castellana, 261,
28046 Madrid, Spain

Principal Investigator:
Dr. Cristina Calvo Rey


Hospital HM Puerta del Sur

Avenida Carlos V, 70,
28938 Móstoles, Madrid, Spain

Principal Investigator:
Dr. Silvina Natalini Martínez


Albera Salut

Carrer Toló, 3, 5,
17491 Peralada, Girona, Spain

Principal Investigator:
Dr. Pyrene Martínez Piera


CAP Centelles
Pla de Mestre, 8,
08540 Centelles, Barcelona, Spain

Principal Investigator:
Dr. Silvia Narejos Pérez


### 3. Eligibility Criteria of the HIPRA-HH-3 trial

Inclusion criteria

Participants must meet all the following criteria to be considered eligible for the study:

1. Adolescents aged from 12 years to less than 18 years at screening.
2. Participant's parent(s)/legal guardian(s) willing and able to sign the informed consent and comply with all study visits and procedures. A written assent was required for all participants in the study.
   Note: Participants were expected to be available for the duration of the study and whose parent(s)/legal guardian could be contacted by telephone during study participation.
3. Participants had received two previous doses of BNT162b2, last dose being at least 6 months before screening.
4. Participants had a body mass index at or above the third percentile according to local child growth standards at screening visit.
5. Healthy participants and participants with pre-existing, chronic, and stable diseases (non-immunocompromised), if these were stable and well-controlled according to the investigator judgment, were eligible for inclusion in the study.
   Note: Healthy participants were determined by medical history, physical examination, and clinical judgment of the investigator. Healthy participants with pre-existing stable diseases, were defined as diseases not requiring significant change in the therapy or hospitalization for worsening disease during the 6 weeks before enrollment.
6. Had a negative Rapid Antigen Test (RAT) at baseline (day 0) before BIMERVAX® vaccine administration.
7. Participants biologically able to have children could be enrolled in the study if the participant fulfilled all the following criteria:
   - Had a negative urine pregnancy test at Screening (day 0), only if biologically able to become pregnant.
   - Had practiced adequate contraception or had abstained from all activities that could result in pregnancy for at least 28 days prior to the booster dose (only if biologically able to become pregnant).
   - Had agreed to continue adequate contraception or abstinence through 3 months following the booster dose.
     - Participants with female reproductive system:
       i. Hormonal contraception (progestogen-only or combined: oral, injectable or transdermal (patch).
       ii. Intrauterine device.
       iii. Vasectomized partner (the vasectomized partner should be the sole partner for that participant).

iv. Condom.

- Participants with male reproductive system:
  i. Vasectomized participants.
  ii. Agreed to use a condom in partners biologically able to become pregnant.

8. Participant had to have a body weight >50 kg at Screening visit to be eligible for the cellular immunology assays.

Exclusion criteria

Participants who met any of the following criteria were excluded from participating in this study:

9. Acute illness with fever ≥ 38.0°C at Screening or within 24 hours prior to vaccination. Participants could be rescheduled for Screening when they had completed 24 hours without fever. Afebrile participants with minor illnesses could be enrolled at the discretion of the investigator.

10. Received medications intended to prevent or treat coronavirus disease 2019 (COVID-19) before Screening, except for BNT162b2 vaccines.

11. Previous or current diagnosis of multisystem inflammatory syndrome in children (MIS-C).

12. Other medical or psychiatric conditions, including recent (within the past year) or active suicidal ideation/behavior or laboratory abnormality that may have increased the risk of study participation or, in the investigator's judgment, made the participant inappropriate for the study.

    Note: This included both conditions that may increase the risk associated with study intervention administration or a condition that may interfere with the interpretation of study results.

13. History of severe adverse reaction associated with a vaccine and/or severe allergic reaction (e.g. anaphylaxis) to any component of the study intervention(s).

14. Immunocompromised individuals defined as those with primary and secondary immune deficiencies and those receiving chemotherapy or immunosuppressant drugs other than steroids and glucocorticoids (maximum 1 mg/kg/day of prednisone or total dose of 20 mg/day by any administration route for a maximum of 30 consecutive days), within 90 days prior to vaccination or during the study.

15. Bleeding diathesis or condition associated with prolonged bleeding that would, in the opinion of the investigator, contraindicate intramuscular injection.

16. Female who was pregnant or breastfeeding.

17. Receipt of blood/plasma products, immunoglobulin, monoclonal antibodies, or receipt of any passive antibody therapy, within 90 days prior to vaccination or during the study.

18. Participation in other studies involving study intervention within 28 days prior to Screening and/or during study participation.

19. Received any non-study vaccine (including seasonal Influenza vaccine) within 14 days before or after Screening. For live or attenuated vaccines, 4 weeks before or after Screening.
20. Had history of illegal substance use or alcohol abuse within the past 2 years.
21. Had history of a diagnosis or other conditions that, in the judgment of the investigator, may affect study endpoint assessment or compromise participant's safety.
22. Individuals who were family members of the Investigators.
23. Individuals with documented medical history of microbiologically confirmed COVID-19 were not eligible for the immunogenicity group.

### 4. Data Collection and Interventions

Data collection

Screening and baseline visits were allowed on the same day (day 0). Medical history, demographics, SARS-CoV-2 rapid antigen test results, and other clinical assessments were recorded at entry. Race and ethnicity assignments were based on self-reports provided by participants using the recommended categories for clinical trials[1]. At screening, participants received diaries to document solicited adverse events (AEs) for seven days following vaccination. Safety assessments were conducted during an in-person visit at day 14 and via telephone at days 7 and 28, with additional in-person visits scheduled as needed.

Treatment-emergent adverse events (TEAEs), serious adverse events (SAEs), medically attended adverse events (MAAEs), and adverse events of special interest (AESIs) through day 28 were also documented. Adverse events were coded using the Medical Dictionary for Regulatory Activities (MedDRA) version 27.0, and intensity was graded according to the National Cancer Institute Common Terminology Criteria for Adverse Events (NCI CTCAE), version 5.0.

Interventions

The first three participants were vaccinated with PHH-1V at least one hour apart and were monitored for one hour. Safety data were collected by phone over 72 hours. If no grade 3 or 4 AEs occurred, vaccination proceeded as scheduled for the remaining participants. Anticoagulants, immunosuppressants, or other immune-modifying agents were prohibited from 90 days pre-day 0 through the study.

### 5. Immunogenicity Assays

#### Humoral immunogenicity

Neutralizing antibody titers against SARS-CoV-2 variants were measured using the inhibitory concentration 50 ($IC_{50}$), defined as the serum dilution required to reduce viral activity by 50%. Measurements were conducted at HIPRA SCIENTIFIC, S.L.U. (Aiguaviva, Spain) using a pseudovirion-based neutralization assay (PBNA). The luciferase-reporter pseudoviruses were pseudotyped with the SARS-CoV-2 spike (S) protein and produced following previously established protocols[2,3].

For the neutralization assay, the pseudovirus stock was first titrated to determine the optimal infectious dose. For this purpose, pseudovirus infectivity was tested by doing serial dilutions and measuring the luminescence obtained in each one. The optimal dilution was that in which a Relative Light Unit (RLU) of approximately 20,000-25,000 was obtained. Once the optimal dilution of the pseudovirus was established, diluted pseudovirus supernatant was preincubated with serial dilutions of the heat-inactivated serum samples, starting at a 1:20 dilution (lower limit of detection), for 1 hour at 37 °C and then added onto ACE-2 overexpressing HEK293T cells. After 48h of incubation, the cells were lysed using Britelite™ Plus luciferase substrate (PerkinElmer, Waltham, MA, USA) and luminescence was recorded over 0.2 seconds using an EnSight™ multimode plate reader (PerkinElmer). Serum neutralization capacity was determined by comparing the RLUs of infected cells treated with each serum dilution to the maximal RLUs (from untreated infected controls) and minimal RLUs (from uninfected cells). The neutralization percentage was calculated using the following formula:

$$\text{Neutralization (\%)} = (RLU_{max} - RLU_{experimental}) / (RLU_{max} - RLU_{min}) \times 100$$

Half-maximal $IC_{50}$ values were derived by plotting the neutralization percentages against the logarithm of serum dilutions and fitting the data to a four-parameter logistic curve using Prism™, version 9.0.2 (GraphPad Software, San Diego, CA, USA).

Total binding antibody titers against the S glycoprotein receptor-binding domain (RBD) of SARS-CoV-2 were analyzed by electrochemiluminescence immunoassay (ECLIA) (Elecsys® Anti-SARS-CoV-2 S, Roche Diagnostics, Rotkreuz, Switzerland), in accordance with previous reports[4].

The percentage of participants achieving a ≥ 4-fold change in total RBD-binding antibodies was based on the post-baseline titer/baseline titer ratio, in accordance with previous reports[4].

#### Cellular immunogenicity

The magnitude of interferon-gamma (IFN-γ)-mediated T-cell response after the administration of PHH-1V booster vaccine was assessed by enzyme-linked immune absorbent spot (ELISpot). The proportion of $CD4^+$ and $CD8^+$ T cells secreting IFN-γ, interleukin (IL)-2, and IL-4 (i.e., Th1/Th2 pathways) were assessed by intracellular cytokine staining (ICS) and flow cytometry.

For both ELISpot and ICS, cryopreserved peripheral blood mononuclear cells (PBMCs) from study participants were thawed in 20% fetal bovine serum (FBS) RPMI (R20) medium and then washed two times with 10% FBS RPMI (R10). For ELISpot, cells were seeded in a 96-well round-bottom plates at $0.2x10^6$ cells/well. Then, PBMCs were restimulated *in vitro* using 15-mer overlapping peptide pools covering the RBD region of the Omicron BA.1 and B.1.351 (Beta) SARS-CoV-2 variants, specified below[4]:

- RBD_BA.1: 84 overlapping peptides spanning the RBD region of the SARS-CoV-2 Omicron BA.1 variant.
- RBD_B.1.351: 84 overlapping peptides spanning the RBD region of the SARS-CoV-2 Beta variant.

Stimulation with individual peptide pools was performed at a final concentration of 2.5 µg/mL. As positive controls, we used stimulation with phytohemagglutinin (PHA) and a cytomegalovirus, Epstein-Barr virus, and influenza virus (CEF) peptide pool (containing 23 MHC class I-restricted T-cell epitopes) at a final concentration of 2.5 µg/mL for each individual peptide in the pool.

Following overnight incubation, ELISpot plates were washed six times with PBS. Spot detection was performed using a two-step antibody-based method: a 1-hour room temperature incubation with biotinylated anti-human IFN-γ antibody, followed by six PBS washes, and another 1-hour incubation with streptavidin-conjugated enzyme. After this, wells were developed with substrate solution, treated with 0.05% Tween 20 in PBS for 10 minutes at room temperature, and washed six times with tap water. Plates were then air-dried upside down, and spots were quantified using the CTL® reader system (Cellular Technology Limited, Cleveland, OH, USA).

Simultaneously, ICS was conducted on PBMCs stimulated with the peptide pools described above. Cells were incubated for 6 hours in the presence of monoclonal antibodies against human CD28 and CD49d (2 µg/mL; BD Pharmingen™, BD Biosciences, San Jose, CA, USA). During the final 4 hours, GolgiPlug™ (contains Brefeldin A; BD Biosciences) was added to inhibit cytokine secretion. After stimulation, cells were washed with PBS supplemented with 0.5% BSA and 0.1% sodium azide, followed by a 20-minute incubation with FcR Blocking Reagent (Miltenyi Biotec, Bergisch Gladbach, Germany). Cells were then stained for 25min with a LIVE/DEAD discrimination probe (LIVE/DEAD™ Fixable Near-IR Dead Cell Stain Kit, Thermo Fisher Scientific, Waltham, MA, USA) and surface markers: CD3 (PerCP), CD4 (BV421), and CD8 (BV510) (BD Biosciences). After two washes, cells were fixed and permeabilized using a Fixation/Permeabilization Solution kit (BD Biosciences) for intracellular staining. Subsequently, cells were re-incubated with FcR Blocking Reagent (Miltenyi Biotec) for 25min, washed, and stained with anti-human cytokine antibodies: IFN-γ (conjugated to allophycocyanin (APC)), IL-2 (phycoerythrin (PE)), and IL-4 (PE-Cy7 tandem) (BD Biosciences). Final washes were performed with Perm/Wash™ buffer (BD Biosciences), and cells were fixed in 1% formaldehyde. Background-subtracted cytokine responses were acquired using a BD FACSCanto™ II flow cytometer (BD Biosciences) and analyzed with FlowJo™ version 10 (Tree Star, Ashland, OR, USA).

<u>Complementary analyses</u>

Beyond the study's primary and secondary endpoints, further subanalyses examined immunogenicity changes against more recently emerged SARS-CoV-2 variants (i.e., Omicron XBB.1.15 and XBB.1.16) by PBNA.

Additionally, immunobridging comparisons of neutralizing and total antibody titers, as well as T-cell responses in adolescents, were performed against those of an external young adult control group from the previous HIPRA-HH-2 study[4].

RBD-specific T-cell responses were also assessed upon *in vitro* restimulation of fresh whole blood samples from participants with a SARS-CoV-2 S protein peptide pool (POOL ONE of XTACT™ SCV2 T Activation kit, Hyris, Milan, Italy) and quantifying *CXCL10* mRNA by RT-qPCR (bKIT™ Immunofinder dqTACT MS, Hyris), following the manufacturer's instructions. *CXCL10* gene, induced by IFN-γ release from activated antigen-specific T cells, serves as a readout of T-cell reactivity against SARS-CoV-2 viral antigens[5]. This complementary technique was included to anticipate potential feasibility of analyzing T-cell responses from smaller blood volumes, particularly in younger children.

Finally, vaccination responders based on T-cell reactivity were defined as participants with ≥ 50 spot-forming units (SFUs) per million PBMCs (SFUs/$10^6$ PBMCs) at day 14 and a ≥ 2-fold increase in SFUs/$10^6$ PBMCs from day 0, as measured by ELISpot and consistent with previous reports[6].

### 6. Statistical Methods

Sample size

An immunobridging analysis compared the geometric mean titer (GMT) in adolescents (≥ 12 to < 18 years) with young adults (aged ≥ 18 to ≤ 25 years) from the previous HIPRA-HH-2 study[4], using a 1.5-fold non-inferiority margin. The young adult dataset included 81 subjects assessed 14 days post-PHH-1V booster.

Of 240 participants vaccinated, immunogenicity was assessed in 88 adolescents (immunogenicity population; IGP), providing an expected statistical power of 81.5% to detect non-inferiority at one-sided 2.5% significance level and assuming a standard deviation (SD)log = 0.40. This exceeded the 80% threshold required for reliable immunogenicity assessment of PHH-1V in adolescents, and within the 80-90% recommended by the guideline ICH E9[7].

Statistical Analyses

The safety population (SP) included all participants receiving a single heterologous PHH-1V booster dose at least 6 months after their second BNT162b2 dose. The IGP comprised participants from SP without documented history of SARS-CoV-2 infection at screening (Table 1). The per-protocol (PP) population is not yet defined, as the trial remains ongoing.

Categorical variables for participant characteristics and safety results are presented as numbers and percentages, while continuous variables use means and corresponding SD. Humoral immunogenicity is expressed as GMT and geometric mean fold rise (GMFR) (day 14/day 0 antibody titers), calculated from log-transformed titers and back-transformed post-summary.

Mixed-effects models for repeated measures (MMRMs) were fitted on $\log_{10}$-transformed neutralizing and binding antibody titers, with fixed effects for visit (day 0 and day 14), age group (adolescents; young adults), and age group-by-visit interaction. Adjusted back-transformed GMT, GMT ratios (young adults vs. adolescents), and 95% confidence intervals (CIs) are reported together with p-values for the age group difference contrasts. Non-inferiority was confirmed if the upper bound of the two-sided 95% CI for GMT ratio against Omicron BA.1 was below the predefined margin of 1.5, per the Food and Drug Administration (FDA) guidance[8]. For XBB.1.5 and XBB.1.16 variants, the same model was fitted excluding the age group fixed effect and interaction. Therefore, adjusted back-transformed GMT, GMT ratio (day 14 vs. day 0), and 95% CIs are reported together with p-values for the visit difference contrasts. GMFR contrasts between age groups were performed using Student's t-tests on $\log_{10}$-transformed data. Furthermore, the percentage of participants exhibiting a ≥ 4-fold increase in binding antibody titers between day 0 and day 14 was determined, and exact Clopper-Pearson CIs were calculated.

T-cell responses (ELISpot) were reported as mean SFU/$10^6$ PBMCs upon stimulation with each variant peptide pool, and cytokine-producing T cells (ICS) were reported as percentage of positive cells. After background subtraction, square root (ELISpot) and arcsine-square root transformations of mean values were employed for quantitative analyses. T-cell responses (using whole blood samples) were reported as *CXCL10* mRNA relative expression. Paired-samples t-tests compared cellular immunity responses between days 0 and 14 among adolescents, and a MMRM was fitted to assess differences between adolescents and young adults for ELISpot data upon Beta stimulation.

Missing data were not imputed. Raw data falling below or above the detection limit of the corresponding assay were imputed as the lower or upper limit of detection, respectively. Statistical analyses were conducted using SAS (version 9.4) and R (version 4.1.3 or later), with a significance threshold for p-values set at 0.05.

**Supplementary Table 1**

Baseline characteristics of study participants included in the subanalysis of humoral immunogenicity (PBNA) against XBB variants and in all T-cell response analyses (ELISpot and ICS) against Omicron BA.1 and Beta variants.

| | **Adolescents (≥ 12 to < 18 years)**[a] (N = 88) | | **Young adults (≥ 18 to ≤ 25 years)**[b] (N = 81) |
|---|---|---|---|
| | PBNA against XBB.1.5 and XBB.1.16 | T-cell response analyses against Omicron BA.1 and Beta | T-cell response analyses against Omicron BA.1 and Beta |
| Participants, n (%) | 44 (50.0) | 16 (18.2) | 9 (11.1) |
| Age | | | |
| Years, mean (SD) | 14.4 (1.5) | 15.2 (1.4) | 23.3 (1.7) |
| Sex, n (%) | | | |
| Female | 28 (31.8) | 8 (9.1) | 8 (9.9) |
| Male | 16 (18.2) | 8 (9.1) | 1 (1.2) |
| Race, n (%) | | | |
| White | 41 (46.6) | 16 (18.2) | 8 (9.9) |
| American Indian or Alaska native | 2 (2.3) | 0 (0.0) | 0 (0.0) |
| Black or African American | 1 (1.1) | 0 (0.0) | 0 (0.0) |
| Asian | 0 (0.0) | 0 (0.0) | 1 (1.2) |
| Ethnicity | | | |
| Hispanic or Latino | 4 (4.5) | 2 (2.3) | 3 (3.7) |
| Not Hispanic or Latino | 40 (45.5) | 14 (15.9) | 6 (7.4) |
| BMI, mean (SD) | 20.7 (2.9) | 21.5 (2.1) | 22.4 (1.7) |
| COVID-19 history | | | |
| Participants with prior COVID-19 episodes, n (%)[c] | 0 (0.0) | 0 (0.0) | 0 (0.0) |
| Time between second dose and booster | | | |
| Months, mean (SD)[d] | 21.9 (3.8) | 24 (2.7) | 10.2 (3.6) |

BMI, body mass index; COVID-19, coronavirus disease 2019; n, number of participants meeting the criterion; N, number of participants in the study population; PBNA, pseudovirion-based neutralization assay; SD, standard deviation.

Percentages have been calculated as n/N*100.

[a]Participants aged ≥ 12 to < 18 years from the HIPRA-HH-3 study.

[b]External control group. Participants aged ≥ 18 to ≤ 25 years from the HIPRA-HH-2 study (NCT05142553)[4].

[c]Documented medical history of microbiologically confirmed COVID-19.

[d]Time between vaccination doses in months is derived as (Date of study booster dose – Date of last booster dose + 1)/30.44 days.

**Supplementary Table 2**

Analysis of neutralizing and total binding antibodies against SARS-CoV-2 at days 0 and 14 after vaccination booster.

| | **Adolescents (≥ 12 to < 18 years)**[a] (N = 88) | | **Young adults (≥ 18 to ≤ 25 years)**[b] (N = 81) | |
|---|---|---|---|---|
| | Day 0 | Day 14 | Day 0 | Day 14 |
| Neutralizing antibodies | | | | |
| *Omicron BA.1* | | | | |
| n (%) | 88 (100.0) | 88 (100.0) | 81 (100.0) | 80 (98.8) |
| GMT | 1303.54 [1016.05; 1672.39] | 24081.34 [19741.36; 29375.43] | 48.48 [37.39; 62.86] | 2340.57 [1900.42; 2882.66] |
| GMT ratio | | | 0.04 [0.03, 0.05]; p < 0.001 | 0.10 [0.07, 0.13]; p < 0.001 |
| GMFR | | 18.47 [14.41; 23.69] | | 47.87 [35.16; 65.18] |
| GMFR ratio | | | | 2.59 [1.75, 3.83]; p < 0.001 |
| *WH1* | | | | |
| n (%) | 88 (100.0) | 88 (100.0) | 81 (100.0) | 80 (98.8) |
| GMT | 780.09 [641.73; 948.28] | 9674.93 [8104.12; 11550.20] | 113.03 [92.22; 138.54] | 2283.44 [1896.33; 2749.58] |
| GMT ratio | | | 0.14 [0.11; 0.19]; p < 0.001 | 0.24 [0.18; 0.31]; p < 0.001 |
| GMFR | | 12.40 [9.90; 15.55] | | 19.98 [15.52; 25.72] |
| GMFR ratio | | | | 1.61 [1.15; 2.25]; p = 0.006 |
| *Beta* | | | | |
| n (%) | 88 (100.0) | 88 (100.0) | 81 (100.0) | 80 (98.8) |
| GMT | 1874.67 [1458.82; 2409.07] | 16037.66 [13136.20; 19579.97] | 98.26 [75.66; 127.62] | 5562.91 [4512.64; 6857.61] |

| | Adolescents (≥ 12 to < 18 years)[a] (N = 88) | | Young adults (≥ 18 to ≤ 25 years)[b] (N = 81) | |
|---|---|---|---|---|
| | Day 0 | Day 14 | Day 0 | Day 14 |
| GMT ratio | | | 0.05 [0.04; 0.07]; p < 0.001 | 0.35 [0.26; 0.46]; p < 0.001 |
| GMFR | | 8.55 [6.76; 10.83] | | 56.12 [40.00; 78.72] |
| GMFR ratio | | | | 6.56 [4.35; 9.88]; p < 0.001 |
| *Delta* | | | | |
| n (%) | 88 (100.0) | 88 (100.0) | 81 (100.0) | 80 (98.8) |
| GMT | 694.11 [550.21; 875.66] | 9781.04 [8217.07; 11642.67] | 64.55 [50.66; 82.23] | 1649.95 [1374.40; 1980.75] |
| GMT ratio | | | 0.09 [0.07; 0.13]; p < 0.001 | 0.17 [0.13; 0.22]; p < 0.001 |
| GMFR | | 14.09 [10.92; 18.18] | | 25.53 [18.75; 34.75] |
| GMFR ratio | | | | 1.81 [1.21; 2.69]; p = 0.003 |
| *XBB.1.5* | | | | |
| n (%) | 44 (50.0) | 44 (50.0) | 0 (0.0) | 0 (0.0) |
| GMT | 92.61 [71.16; 120.54] | 1031.94 [792.86; 1343.11] | | |
| GMT ratio | | 11.14 [8.25; 15.05]; p < 0.001 | | |
| *XBB.1.16* | | | | |
| n (%) | 44 (50.0) | 44 (50.0) | 0 (0.0) | 0 (0.0) |
| GMT | 150.04 [116.20; 193.74] | 1447.99 [1121.38; 1869.72] | | |
| GMT ratio | | 9.65 [7.27; 12.80]; p < 0.001 | | |
| Binding antibodies | | | | |
| n (%) | 87 (98.9) | 88 (100.0) | 81 (100.0) | 79 (97.5) |
| GMT | 7960.79 [6398.63; 9904.35] | 98038.63 [84938.91; 113158.65] | 854.88 [681.60; 1072.20] | 43565.17 [37451.68; 50676.61] |
| GMT ratio | | | 0.11 [0.08; 0.15]; p < 0.001 | 0.44 [0.36; 0.55]; p < 0.001 |

| | Adolescents (≥ 12 to < 18 years)[a] (N = 88) | | Young adults (≥ 18 to ≤ 25 years)[b] (N = 81) | |
|---|---|---|---|---|
| | Day 0 | Day 14 | Day 0 | Day 14 |
| n (%) | | 87 (98.9) | | 79 (97.5) |
| GMFR | | 12.39 [9.80; 15.67] | | 51.08 [41.11, 63.47] |
| GMFR ratio | | | | 4.12 [3.00; 5.67]; $p < 0.001$ |

CI, confidence interval; GMT, geometric mean titer; GMFR, geometric mean fold rise. n, subjects with available data included in the analysis; MMRM, mixed-effects model for repeated measures; N, subjects included in the study population.

[a]Participants aged ≥ 12 to < 18 years from the HIPRA-HH-3 study.

[b]External control group. Participants aged ≥ 18 to ≤ 25 years from the HIPRA-HH-2 study (NCT05142553)[4].

Percentages have been calculated as n/N*100.

GMT is shown as back-transformed adjusted group mean [95% CI]; GMT ratio is shown as age group ratio for adjusted GMT (young adults vs. adolescents) [95% CI] followed by p-value for ratio = 1. MMRM was fitted to assess the endpoints on the log10 scale.

For variants XBB.1.5 and XBB.1.16, GMT is shown as back-transformed adjusted group mean [95% CI]; GMT ratio is shown as visit ratio for adjusted GMT (day 14 vs day 0) [95% CI] followed by p-value for ratio = 1. MMRM was fitted to assess the endpoints on the log10 scale.

GMFR is shown as back-transformed fold rise of group means between visits [95% CI]; GMFR ratio is shown as age group ratio for GMFR (young adults vs. adolescents) [95% CI] followed by p-value for ratio = 1. Student's t-test was performed to assess the endpoints on the log10 scale.

**Supplementary Table 3**

TEAEs through day 28. Data are shown as number of events as well as the number of participants and percentage in relation to the safety population (N = 240). If a participant experienced more than one event, the participant is counted once for each SOC and once for each PT.

| | Events, n | Participants, n (%) |
|---|---|---|
| **TEAEs** | | |
| Total TEAEs | 865 | 206 (85.8) |
| **TEAEs by SOC / PT** | | |
| General disorders and administration site conditions | 558 | 195 (81.3) |
| Injection site pain | 313 | 187 (77.9) |
| Fatigue | 82 | 73 (30.4) |
| Malaise | 75 | 67 (27.9) |
| Axillary pain | 29 | 28 (11.7) |
| Injection site induration | 26 | 26 (10.8) |
| Injection site erythema | 16 | 16 (6.7) |
| Pyrexia | 9 | 9 (3.8) |
| Discomfort | 3 | 3 (1.3) |
| Vessel puncture site pain | 2 | 2 (0.8) |
| Asthenia | 1 | 1 (0.4) |
| Injection site swelling | 1 | 1 (0.4) |
| Secretion discharge | 1 | 1 (0.4) |
| Nervous system disorders | 98 | 79 (32.9) |
| Headache | 92 | 77 (32.1) |
| Presyncope | 2 | 2 (0.8) |
| Syncope | 2 | 2 (0.8) |
| Dizziness | 1 | 1 (0.4) |
| Migraine | 1 | 1 (0.4) |
| Musculoskeletal and connective tissue disorders | 82 | 52 (21.7) |
| Myalgia | 41 | 39 (16.3) |
| Arthralgia | 39 | 35 (14.6) |
| Pain in extremity | 1 | 1 (0.4) |
| Synovial cyst | 1 | 1 (0.4) |
| Gastrointestinal disorders | 44 | 33 (13.8) |
| Diarrhea | 21 | 18 (7.5) |
| Vomiting | 14 | 14 (5.8) |
| Nausea | 6 | 6 (2.5) |
| Odynophagia | 2 | 2 (0.8) |
| Abdominal pain | 1 | 1 (0.4) |
| Infections and infestations | 23 | 22 (9.2) |
| Pharyngitis | 5 | 5 (2.1) |
| Nasopharyngitis | 3 | 3 (1.3) |

| | Events, n | Participants, n (%) |
|---|---|---|
| Viral infection | 3 | 3 (1.3) |
| Upper respiratory tract infection | 2 | 2 (0.8) |
| Bronchitis | 1 | 1 (0.4) |
| Conjunctivitis | 1 | 1 (0.4) |
| Gastroenteritis | 1 | 1 (0.4) |
| Otitis externa | 1 | 1 (0.4) |
| Pharyngotonsillitis | 1 | 1 (0.4) |
| Pneumonia | 1 | 1 (0.4) |
| Postoperative wound infection | 1 | 1 (0.4) |
| Pustule | 1 | 1 (0.4) |
| Rhinitis | 1 | 1 (0.4) |
| Tonsillitis bacterial | 1 | 1 (0.4) |
| Blood and lymphatic system disorders | 23 | 21 (8.8) |
| Lymphadenopathy | 19 | 19 (7.9) |
| Anemia | 1 | 1 (0.4) |
| Mesenteric lymphadenitis | 1 | 1 (0.4) |
| Neutropenia | 1 | 1 (0.4) |
| Splenomegaly | 1 | 1 (0.4) |
| Respiratory, thoracic and mediastinal disorders | 12 | 12 (5.0) |
| Cough | 5 | 5 (2.1) |
| Bronchospasm | 2 | 2 (0.8) |
| Oropharyngeal pain | 2 | 2 (0.8) |
| Asthma | 1 | 1 (0.4) |
| Epistaxis | 1 | 1 (0.4) |
| Rhinitis allergic | 1 | 1 (0.4) |
| Reproductive system and breast disorders | 7 | 6 (2.5) |
| Premenstrual pain | 5 | 5 (2.1) |
| Intermenstrual bleeding | 1 | 1 (0.4) |
| Menstruation irregular | 1 | 1 (0.4) |
| Skin and subcutaneous tissue disorders | 6 | 6 (2.5) |
| Erythema | 4 | 4 (1.7) |
| Hyperhidrosis | 1 | 1 (0.4) |
| Rash | 1 | 1 (0.4) |
| Injury, poisoning and procedural complications | 3 | 3 (1.3) |
| Ligament sprain | 2 | 2 (0.8) |
| Craniocerebral injury | 1 | 1 (0.4) |
| Investigations | 3 | 3 (1.3) |
| Blood bilirubin increased | 2 | 2 (0.8) |
| Neutrophil count increased | 1 | 1 (0.4) |
| Congenital, familial and genetic disorders | 2 | 2 (0.8) |

| | Events, n | Participants, n (%) |
|---|---|---|
| Congenital scoliosis | 1 | 1 (0.4) |
| Gilberts syndrome | 1 | 1 (0.4) |
| Psychiatric disorders | 2 | 2 (0.8) |
| Anxiety | 1 | 1 (0.4) |
| Insomnia | 1 | 1 (0.4) |
| Ear and labyrinth disorders | 1 | 1 (0.4) |
| Ear pain | 1 | 1 (0.4) |
| Social circumstances | 1 | 1 (0.4) |
| Menarche | 1 | 1 (0.4) |
| **TEAEs by intensity** | | |
| Mild | 733 | 151 (62.9) |
| Moderate | 113 | 47 (19.6) |
| Severe | 19 | 8 (3.3) |
| **TEAEs by relationship to study treatment** | | |
| Related | 750 | 198 (82.5) |
| Unrelated | 115 | 8 (3.3) |
| **Treatment-emergent MAAEs** | | |
| Related | 0 | 0 (0.0) |
| Unrelated | 120 | 66 (27.5) |
| **Treatment-emergent AESIs** | | |
| Total treatment-emergent AESIs | 0 | 0 (0.0) |
| **Treatment-emergent SAEs** | | |
| Total treatment-emergent SAEs | 0 | 0 (0.0) |

AESIs, adverse events of special interest; MAAEs, medically attended adverse events; PT, preferred term; SAEs, serious adverse events; SOC, system organ class; TEAEs, treatment-emergent adverse events.

TEAEs were defined as any AE with onset on or after the administration of study treatment through day 28 or any AE that was present at day 0 but worsened in intensity or was subsequently considered drug-related by the Investigator through the end of the study.

**Supplementary Table 4**

Solicited local reactions through day 7. Data are shown as number of events as well as the number of participants and percentage in relation to the safety population (N = 240). If a participant experienced more than one event, the participant is counted once for each type of event.

| | Events (n) | Participants, n (%) |
|---|---|---|
| *Cumulative events from day 0 to day 7* | | |
| Total events | 758 | 189 (78.8) |
| Pain | 365 | 174 (72.5) |
| Tenderness | 310 | 134 (55.8) |
| Induration/swelling | 50 | 25 (10.4) |
| Erythema/redness | 33 | 19 (7.9) |
| *Day 0* | | |
| Total number of events | 280 | 158 (65.8) |
| Pain | 139 | 139 (57.9) |
| Tenderness | 117 | 117 (48.8) |
| Induration/swelling | 17 | 17 (7.1) |
| Erythema/redness | 7 | 7 (2.9) |
| *Day 1* | | |
| Total number of events | 280 | 160 (66.7) |
| Pain | 144 | 144 (60.0) |
| Tenderness | 105 | 105 (43.8) |
| Induration/swelling | 16 | 16 (6.7) |
| Erythema/redness | 15 | 15 (6.3) |
| *Day 2* | | |
| Total number of events | 111 | 71 (29.6) |
| Pain | 49 | 49 (20.4) |
| Tenderness | 48 | 48 (20.0) |
| Induration/swelling | 9 | 9 (3.8) |
| Erythema/redness | 5 | 5 (2.1) |
| *Day 3* | | |
| Total number of events | 43 | 29 (12.1) |
| Pain | 18 | 18 (7.5) |
| Tenderness | 19 | 19 (7.9) |
| Induration/swelling | 4 | 4 (1.7) |
| Erythema/redness | 2 | 2 (0.8) |
| *Day 4* | | |
| Total number of events | 18 | 12 (5.0) |
| Pain | 7 | 7 (2.9) |
| Tenderness | 8 | 8 (3.3) |
| Induration/swelling | 2 | 2 (0.8) |
| Erythema/redness | 1 | 1 (0.4) |
| *Day 5* | | |
| Total number of events | 10 | 5 (2.1) |

| | Events (n) | Participants, n (%) |
|---|---|---|
| Pain | 3 | 3 (1.3) |
| Tenderness | 5 | 5 (2.1) |
| Induration/swelling | 1 | 1 (0.4) |
| Erythema/redness | 1 | 1 (0.4) |
| *Day 6* | | |
| Total number of events | 11 | 5 (2.1) |
| Pain | 3 | 3 (1.3) |
| Tenderness | 5 | 5 (2.1) |
| Induration/swelling | 1 | 1 (0.4) |
| Erythema/redness | 2 | 2 (0.8) |
| *Day 7* | | |
| Total number of events | 5 | 4 (1.7) |
| Pain | 2 | 2 (0.8) |
| Tenderness | 3 | 3 (1.3) |
| Induration/swelling | 0 | 0 (0.0) |
| Erythema/redness | 0 | 0 (0.0) |

**Supplementary Table 5**

Solicited systemic AEs through day 7. Data are shown as number of events as well as the number of participants and percentage in relation to the safety population (N = 240). If a participant experienced more than one event, the participant is counted once for each type of event.

| | Events (n) | Participants, n (%) |
|---|---|---|
| *Cumulative events from day 0 to day 7* | | |
| Total AEs | 762 | 136 (56.7) |
| Headache | 163 | 77 (32.1) |
| Malaise | 141 | 70 (29.2) |
| Fatigue | 141 | 73 (30.4) |
| Muscle pain | 82 | 39 (16.3) |
| Joint pain | 63 | 35 (14.6) |
| Axillary pain | 55 | 27 (11.3) |
| Enlarged lymph nodes (lymphadenopathy) | 42 | 17 (7.1) |
| Nausea/vomiting | 28 | 18 (7.5) |
| Fever | 10 | 7 (2.9) |
| Diarrhea | 37 | 17 (7.1) |
| *Day 0* | | |
| Total number of events | 176 | 80 (33.3) |
| Headache | 31 | 31 (12.9) |
| Malaise | 35 | 35 (14.6) |
| Fatigue | 32 | 32 (13.3) |
| Muscle pain | 25 | 25 (10.4) |
| Joint pain | 21 | 21 (8.8) |
| Axillary pain | 11 | 11 (4.6) |
| Enlarged lymph nodes (lymphadenopathy) | 7 | 7 (2.9) |
| Nausea/vomiting | 7 | 7 (2.9) |
| Fever | 4 | 4 (1.7) |
| Diarrhea | 3 | 3 (1.3) |
| *Day 1* | | |
| Total number of events | 242 | 105 (43.8) |
| Headache | 47 | 47 (19.6) |
| Malaise | 48 | 48 (20.0) |
| Fatigue | 49 | 49 (20.4) |
| Muscle pain | 31 | 31 (12.9) |
| Joint pain | 25 | 25 (10.4) |
| Axillary pain | 17 | 17 (7.1) |
| Enlarged lymph nodes (lymphadenopathy) | 11 | 11 (4.6) |
| Nausea/vomiting | 8 | 8 (3.3) |
| Fever | 3 | 3 (1.3) |

| | Events (n) | Participants, n (%) |
|---|---|---|
| Diarrhea | 3 | 3 (1.3) |
| *Day 2* | | |
| Total number of events | 137 | 65 (27.1) |
| Headache | 30 | 30 (12.5) |
| Malaise | 22 | 22 (9.2) |
| Fatigue | 26 | 26 (10.8) |
| Muscle pain | 17 | 17 (7.1) |
| Joint pain | 9 | 9 (3.8) |
| Axillary pain | 16 | 16 (6.7) |
| Enlarged lymph nodes (lymphadenopathy) | 7 | 7 (2.9) |
| Nausea/vomiting | 4 | 4 (1.7) |
| Fever | 0 | 0 (0.0) |
| Diarrhea | 6 | 6 (2.5) |
| *Day 3* | | |
| Total number of events | 58 | 34 (14.2) |
| Headache | 14 | 14 (5.8) |
| Malaise | 8 | 8 (3.3) |
| Fatigue | 12 | 12 (5.0) |
| Muscle pain | 2 | 2 (0.8) |
| Joint pain | 3 | 3 (1.3) |
| Axillary pain | 9 | 9 (3.8) |
| Enlarged lymph nodes (lymphadenopathy) | 5 | 5 (2.1) |
| Nausea/vomiting | 1 | 1 (0.4) |
| Fever | 0 | 0 (0.0) |
| Diarrhea | 4 | 4 (1.7) |
| *Day 4* | | |
| Total number of events | 38 | 22 (9.2) |
| Headache | 10 | 10 (4.2) |
| Malaise | 9 | 9 (3.8) |
| Fatigue | 5 | 5 (2.1) |
| Muscle pain | 1 | 1 (0.4) |
| Joint pain | 1 | 1 (0.4) |
| Axillary pain | 2 | 2 (0.8) |
| Enlarged lymph nodes (lymphadenopathy) | 4 | 4 (1.7) |
| Nausea/vomiting | 1 | 1 (0.4) |
| Fever | 0 | 0 (0.0) |
| Diarrhea | 5 | 5 (2.1) |
| *Day 5* | | |
| Total number of events | 46 | 23 (9.6) |
| Headache | 13 | 13 (5.4) |
| Malaise | 10 | 10 (4.2) |

| | Events (n) | Participants, n (%) |
|---|---|---|
| Fatigue | 6 | 6 (2.5) |
| Muscle pain | 3 | 3 (1.3) |
| Joint pain | 1 | 1 (0.4) |
| Axillary pain | 0 | 0 (0.0) |
| Enlarged lymph nodes (lymphadenopathy) | 4 | 4 (1.7) |
| Nausea/vomiting | 4 | 4 (1.7) |
| Fever | 0 | 0 (0.0) |
| Diarrhea | 5 | 5 (2.1) |
| *Day 6* | | |
| Total number of events | 35 | 19 (7.9) |
| Headache | 8 | 8 (3.3) |
| Malaise | 6 | 6 (2.5) |
| Fatigue | 7 | 7 (2.9) |
| Muscle pain | 2 | 2 (0.8) |
| Joint pain | 2 | 2 (0.8) |
| Axillary pain | 0 | 0 (0.0) |
| Enlarged lymph nodes (lymphadenopathy) | 2 | 2 (0.8) |
| Nausea/vomiting | 1 | 1 (0.4) |
| Fever | 2 | 2 (0.8) |
| Diarrhea | 5 | 5 (2.1) |
| *Day 7* | | |
| Total number of events | 30 | 16 (6.7) |
| Headache | 10 | 10 (4.2) |
| Malaise | 3 | 3 (1.3) |
| Fatigue | 4 | 4 (1.7) |
| Muscle pain | 1 | 1 (0.4) |
| Joint pain | 1 | 1 (0.4) |
| Axillary pain | 0 | 0 (0.0) |
| Enlarged lymph nodes (lymphadenopathy) | 2 | 2 (0.8) |
| Nausea/vomiting | 2 | 2 (0.8) |
| Fever | 1 | 1 (0.4) |
| Diarrhea | 6 | 6 (2.5) |

AEs, adverse events.

**Supplementary Table 6**

Unsolicited local and systemic AEs through day 28. Data are shown as number of events as well as the number of participants and percentage in relation to the safety population (N = 240). If a participant experienced more than one event, the participant is counted once for each type of event. Some events reported as unsolicited occurred within 7 days and could be considered as solicited.

| | **Events, n** | **Participants, n (%)** |
|---|---|---|
| Total AEs | 95 | 62 (25.8) |
| Cough | 5 | 5 (2.1) |
| Pharyngitis | 5 | 5 (2.1) |
| Premenstrual pain | 5 | 5 (2.1) |
| Injection site pain | 4 | 4 (1.7) |
| Nasopharyngitis | 3 | 3 (1.3) |
| Viral infection | 3 | 3 (1.3) |
| Arthralgia | 2 | 2 (0.8) |
| Blood bilirubin increased | 2 | 2 (0.8) |
| Bronchospasm | 2 | 2 (0.8) |
| Injection site induration | 2 | 2 (0.8) |
| Ligament sprain | 2 | 2 (0.8) |
| Lymphadenopathy | 2 | 2 (0.8) |
| Odynophagia | 2 | 2 (0.8) |
| Oropharyngeal pain | 2 | 2 (0.8) |
| Presyncope | 2 | 2 (0.8) |
| Pyrexia | 2 | 2 (0.8) |
| Syncope | 2 | 2 (0.8) |
| Upper respiratory tract infection | 2 | 2 (0.8) |
| Vessel puncture site pain | 2 | 2 (0.8) |
| Vomiting | 2 | 2 (0.8) |
| Abdominal pain | 1 | 1 (0.4) |
| Anemia | 1 | 1 (0.4) |
| Anxiety | 1 | 1 (0.4) |
| Asthenia | 1 | 1 (0.4) |
| Asthma | 1 | 1 (0.4) |
| Axillary pain | 1 | 1 (0.4) |
| Bronchitis | 1 | 1 (0.4) |
| Congenital scoliosis | 1 | 1 (0.4) |
| Conjunctivitis | 1 | 1 (0.4) |
| Craniocerebral injury | 1 | 1 (0.4) |
| Diarrhea | 1 | 1 (0.4) |
| Dizziness | 1 | 1 (0.4) |
| Ear pain | 1 | 1 (0.4) |
| Epistaxis | 1 | 1 (0.4) |
| Erythema | 1 | 1 (0.4) |
| Fatigue | 1 | 1 (0.4) |

| | Events, n | Participants, n (%) |
|---|---|---|
| Gastroenteritis | 1 | 1 (0.4) |
| Gilberts syndrome | 1 | 1 (0.4) |
| Headache | 1 | 1 (0.4) |
| Hyperhidrosis | 1 | 1 (0.4) |
| Injection site erythema | 1 | 1 (0.4) |
| Insomnia | 1 | 1 (0.4) |
| Intermenstrual bleeding | 1 | 1 (0.4) |
| Menarche | 1 | 1 (0.4) |
| Menstruation irregular | 1 | 1 (0.4) |
| Mesenteric lymphadenitis | 1 | 1 (0.4) |
| Migraine | 1 | 1 (0.4) |
| Myalgia | 1 | 1 (0.4) |
| Neutropenia | 1 | 1 (0.4) |
| Neutrophil count increased | 1 | 1 (0.4) |
| Otitis externa | 1 | 1 (0.4) |
| Pain in extremity | 1 | 1 (0.4) |
| Pharyngotonsillitis | 1 | 1 (0.4) |
| Pneumonia | 1 | 1 (0.4) |
| Postoperative wound infection | 1 | 1 (0.4) |
| Pustule | 1 | 1 (0.4) |
| Rash | 1 | 1 (0.4) |
| Rhinitis | 1 | 1 (0.4) |
| Rhinitis allergic | 1 | 1 (0.4) |
| Secretion discharge | 1 | 1 (0.4) |
| Splenomegaly | 1 | 1 (0.4) |
| Tonsillitis bacterial | 1 | 1 (0.4) |

AEs, adverse events.

**Supplementary Figure**

**Analysis of humoral immunogenicity following PHH-1V booster vaccine**. **(A)** Fold rise in neutralizing antibody titers against multiple SARS-CoV-2 variants in adolescents (N = 88) vs. young adults (N = 81) before (day 0) and two weeks after (day 14) receiving the booster immunization. **(B)** Neutralizing antibody titers against XBB.1.5 and XBB.1.16 variants in adolescents at day 0 vs. day 14 post-booster (N = 44). **(C)** Fold rise in neutralizing titers against XBB.1.5 and XBB.1.16 variants (N = 44). **(D)** Fold rise in SARS-CoV-2 binding antibodies in adolescents (N = 87) vs. young adults (N = 81) at days 0 and 14. Young adults (aged 18-25; HIPRA-HH-2 study [Corominas *et al.*[4]]) served as the active control group. CI, confidence interval. Statistical analyses were conducted using a Student's t-test. **p < 0.01; ***p < 0.001.

### References (Supplementary Information)
